# mTOR inhibition improves the formation of functional T cell memory following COVID-19 vaccination of kidney transplant recipients

**DOI:** 10.1101/2023.03.27.23287773

**Authors:** Griffith B. Perkins, Matthew J. Tunbridge, Cheng Sheng Chai, Christopher M. Hope, Arthur Eng Lip Yeow, Tania Salehi, Julian Singer, Bree Shi, Makutiro G. Masavuli, Zelalem Addis Mekonnen, Pablo Garcia-Valtanen, Svjetlana Kireta, Julie K. Johnston, Christopher J. Drogemuller, Beatrice Z. Sim, Shane M. Spencer, Benedetta C. Sallustio, Iain Comerford, George Bouras, Daniela Weiskopf, Alessandro Sette, Anupriya Aggarwal, Vanessa Milogiannakis, Anouschka Akerman, Stuart Turville, Plinio R. Hurtado, Tracey Ying, Pravin Hissaria, Simon C. Barry, Steven J. Chadban, Branka Grubor-Bauk, P. Toby Coates

## Abstract

Inadequate immune response to vaccination is a long-standing problem faced by immunosuppressed kidney transplant recipients (KTRs), requiring novel strategies to improve vaccine efficacy. In this study, the potential of mechanistic target of rapamycin inhibitors (mTORi) to improve T cell responses to COVID-19 vaccination was investigated. Following primary vaccination with adenoviral (ChAdOx1) or mRNA (BNT162b2) COVID-19 vaccines, KTRs receiving rapamycin demonstrated T cell responses greater than those of healthy individuals, characterized by increased frequencies of vaccine-specific central memory, effector memory and T_EMRA_ T cells, in both the CD4^+^ and CD8^+^ compartments. Relative to standard-of-care triple therapy, mTORi-based therapy was associated with a 12-fold greater functional T cell response to primary vaccination of KTRs. The use of rapamycin to augment T cell responses to COVID-19 booster (third dose) vaccination was next investigated in a randomized, controlled trial. Immunosuppression modification with rapamycin was feasible and well-tolerated, but did not improve vaccine-specific T cell responses in this cohort. To understand the parameters for effective use of rapamycin as a vaccine adjuvant, mice were treated with rapamycin before primary or booster vaccination with ancestral and/or Omicron COVID-19 vaccines. Supporting the findings from KTRs, significant enhancement of functional and stem-like memory T cell responses was observed when rapamycin was administered from the time of primary, rather than booster, vaccination. Collectively, a positive effect of mTOR inhibitors on vaccine-induced T cell immunity against COVID-19 in humans was demonstrated.

**One Sentence Summary:** Rapamycin use at the time of primary COVID-19 vaccination augments the formation of functional, vaccine-specific T cell memory in immunosuppressed kidney transplant recipients.

## INTRODUCTION

Kidney transplant recipients (KTRs) are at high risk of severe disease from SARS-CoV-2 infection (*1, 2*). Immunocompromised individuals were prioritized for vaccination in numerous countries, however it is now clear that COVID-19 vaccines are poorly immunogenic in this group, and afford limited real-world protection from severe disease (*3–11*). Vaccine boosters have been found to increase antibody titers in those with poor responses to primary vaccination, but virus neutralizing titers remain suboptimal in the majority of patients after a third or fourth dose, and a subgroup of patients remain seronegative (*10–12*). Indirect approaches to protect KTRs through ring vaccination may be beneficial in the immediate-term (*13*), however a strategy to directly boost immunity in immunosuppressed individuals is urgently needed (*14*). Such a strategy would likely center on transient modification of immunosuppression, and must strike the balance of enhancing vaccine immunogenicity and efficacy without increasing the risk of transplant rejection.

KTRs require lifelong immunosuppression to prevent kidney rejection, and a combination of a calcineurin inhibitor (CNI; e.g. tacrolimus), anti-metabolite (e.g. mycophenolate mofetil), and steroid (e.g. prednisolone) is the current global standard-of-care (SOC). Alternatively, mechanistic target of rapamycin (mTOR) complex 1 (mTORC1) inhibitors, rapamycin (sirolimus) and everolimus, may provide some advantages as an alternative to CNIs, in reducing nephrotoxicity (*15*), or to anti-metabolites in reducing risk of viral infection (*16*).

In preclinical models of vaccination, rapamycin has been found to paradoxically promote the expansion of antigen-specific T cell memory (*17–19*). Treatment of mice or non-human primates with rapamycin over the peri-vaccination period results in an increase in the frequency and functionality of the resultant antigen specific CD8^+^ memory T cell pool (*17, 18*). In mice this effect is mediated by direct inhibition mTORC1 in CD8^+^ T cells, and provides enhanced viral control (*17*). Consistent with this, mTORi use by transplant recipients is associated with reduced viral reactivation and lower incidence of some cancers, although whether these observations result from enhanced T cell memory is yet to be established (*16, 20–22*). Importantly, this augmentation of T cell memory response in mice is selectively observed when T cell antigens are presented in the context of viral infection, and not in the context of transplantation (*23*). As such, rapamycin may be uniquely able to enhance antiviral immunity while suppressing the transplant allo-response.

Here, humoral and cellular immune responses to vaccination against SARS-CoV-2 in healthy individuals and in KTRs receiving alternative mTORi-based immunosuppression regimens were evaluated. We report superior cellular and humoral immune responses to primary COVID-19 vaccination in KTRs receiving mTORi-based immunosuppression compare to the SOC, and present the first in-human evidence for a boosting effect of mTORC1 inhibition on the formation of functional, antigen-specific T cell memory to vaccination. The potential for immunosuppression modification with rapamycin as a strategy to boost the immune response of kidney transplant recipients to ancestral and variant SARS-CoV-2 vaccines was further evaluated in a pre-clinical study and a randomized, controlled trial.

## RESULTS

### mTOR inhibitors are associated with superior induction of humoral and cellular immunity to primary COVID-19 vaccination as compared to calcineurin inhibitors in a three-drug kidney transplant maintenance regimen

Healthy individuals and KTRs were prospectively recruited to receive two doses of BNT162b2 or ChAdOx1 as part of the REVAX trial (ACTRN12621000532808; fig. S1) and in accordance with Australian health guidelines (Fig. 1). KTRs were receiving a variety of immunosuppressive drug regimens (Table 1), and healthy (non-transplanted) controls of comparable age and sex were recruited from amongst patients’ cohabitants (Table 1). Patients and cohabitants were vaccinated concurrently, and peripheral blood samples were collected three weeks (median 22 days, IQR 22-25) after the second vaccine dose for analysis of SARS-CoV-2-specific immunity (Fig. 1).

**Figure 1.**
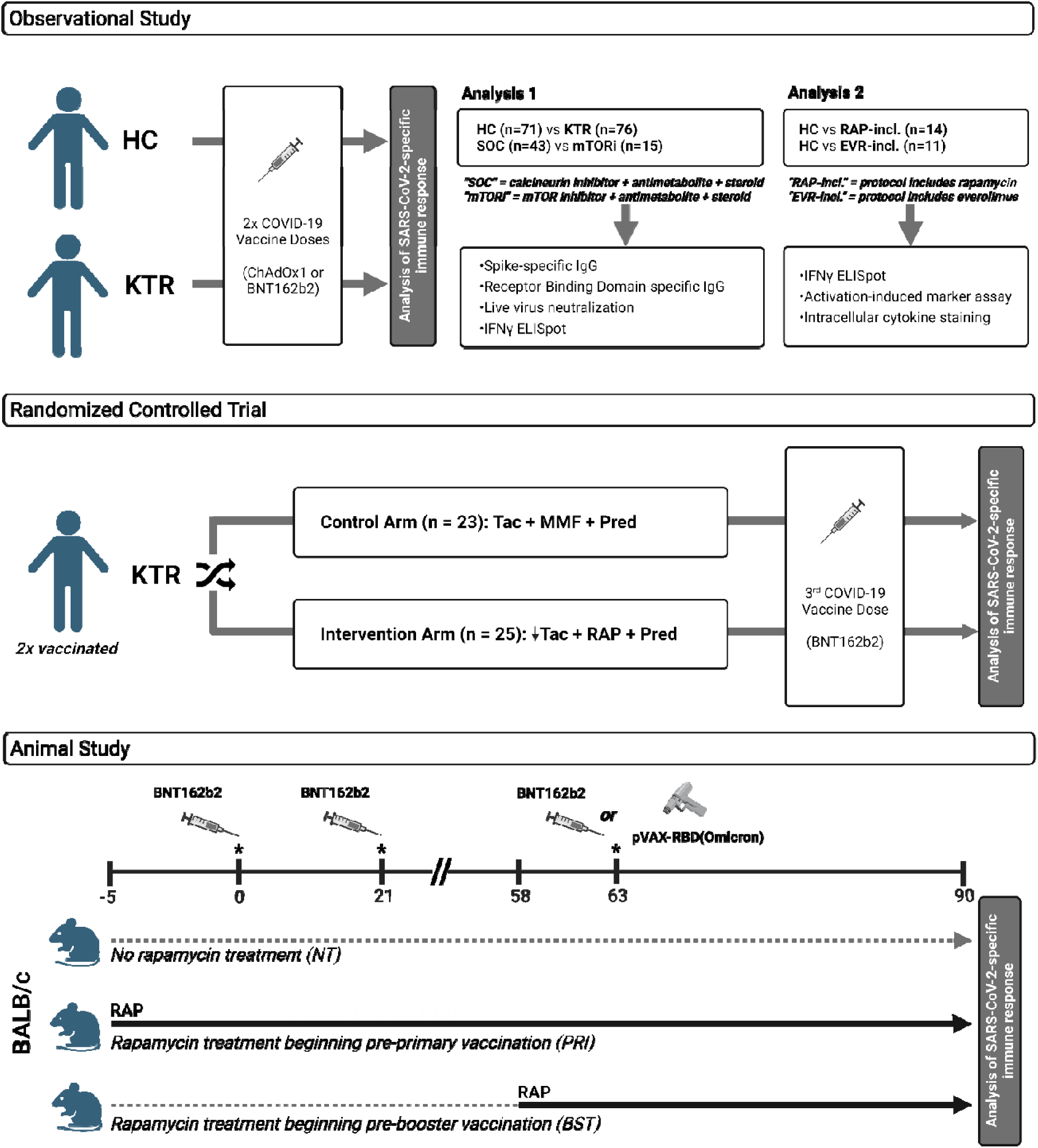
Study design. ***Observational Study:*** Unvaccinated, SARS-CoV-2 infection-naïve kidney transplant recipients (KTR; n = 76) and healthy controls (HC; n = 71) were administered two doses of ChAdOx1 or BNT162b2 vaccine as part of the REVAX trial (ACTRN12621000532808), and peripheral blood samples were collected 21 days post-second dose. A comprehensive analysis of the functional humoral and cellular SARS-CoV-2 Spike-specific immune response was performed. *Analysis 1:* Spike-specific immunity was compared between HCs and KTRs, and between KTRs receiving CNI-based standard-of-care (‘SOC’; n = 48) and mTORi-based (‘mTORi’; n = 15) three-drug regimens. *Analysis 2:* Based on the high IFNγ ELISpot response observed for the mTORi group in Analysis 1, a second analysis was performed comparing Spike-specific T cell responses in KTRs receiving an mTORi (n = 25, independent of immunosuppresion protocol) to that of healthy individuals, in order to test the hypothesis that mTORi use is associated with a boost in the antigen-specific T cell memory response to vaccination. mTORi users were stratified into those receiving rapamycin (n = 14) and everolimus (n = 11), and the phenotype and function of Spike-specific T cells in this subcohort assessed by AIM and ICS assays by flow cytometry. ***Randomized controlled trial:*** a randomized, controlled trial was performed across two academic transplant centers in Australia to measure the effect of immunosuppression modification with rapamycin on humoral and cellular immune response to COVID-19 booster vaccination. KTRs receiving SOC triple therapy were randomized to remain on existing treatment or switch to the intervention arm: cease mycophenolate mofetil, start rapamycin and reduce tacrolimus. Participants received a third COVID-19 vaccine dose (BNT162b2), and cellular and humoral immunity was assessed 4 weeks later. ***Animal study:*** BALB/c mice (n=7 per group) were vaccinated (intra-muscularly (IM)) with two doses of BNT162b2 COVID-19 vaccine (1 μg), boosted with BNT162b2 (1 μg, IM), or with Omicron-RBD DNA vaccine (50 μg, intra-dermally (ID)). Mice were started on rapamycin prior to the primary vaccination course (PRI), or prior to booster vaccination (BST) with the homologous or Omicron variant vaccine, and the effect of rapamycin on humoral and cellular immune responses were evaluated. Created with BioRender.com.

**Table 1.** Participant characteristics – cohort for Analysis 1 (see Fig. 1)

| Characteristic | Healthy cohabitants (n = 71) | KTR (n = 76) |
| --- | --- | --- |
| Age | 59.4 ± 12.2 (μ ± SD) | 59.5 ± 17.0 (μ ± SD) |
| Gender | F: 48 (68%)<br>M: 23 (32%) | F: 24 (32%)<br>M: 52 (68%) |
| Vaccine | ChAdOx1-S: 36 (51%)<br>BNT162b2: 35 (49%) | ChAdOx1-S: 44 (58%)<br>BNT162b2: 32 (42%) |
| Graft number | NA | First graft: 64 (84%)<br>Second graft: 12 (16%) |
| Cause of ESKD |  |  |
| Glomerulonephritis |  | 32 (42%) |
| Other |  | 15 (20%) |
| Polycystic disease | NA | 17 (22%) |
| Renovascular |  | 3 (4%) |
| Diabetic nephropathy |  | 2 (3%) |
| Unknown |  | 7 (9%) |
| Current immunosuppression therapy | NA | CNI-based: 40 (52%)<br>mTORi-based: 25 (30%)<br>Other: 11 (17%) |
| Comorbidities |  |  |
| Hypertension | 24 (34%) | 71 (93%) |
| Diabetes mellitus | 10 (14%) | 27 (36%) |
| IHD | 2 (3%) | 10 (13%) |
| PVD | 0 (0%) | 2 (3%) |
| CerebroVD | 0 (0%) | 1 (1%) |
| Chronic lung disease | 3 (4%) | 5 (7%) |
| Dyslipidaemia | 20 (28%) | 44 (58%) |
| Cancer (inactive) | 4 (6%) | 15 (20%) |
| Kidney donor | 10 (14%) | NA |
| Other | 36 (51%) | 56 (74%) |
| Medications |  |  |
| Antihypertensive | 21 (30%) | 69 (91%) |
| Lipid-lowering | 20 (28%) | 51 (67%) |
| Diabetic therapy | 8 (11%) | 17 (22%) |
| Antiplatelet | 4 (6%) | 20 (26%) |
| Anticoagulant | 2 (3%) | 15 (20%) |
| Anti-arrhythmic | 1 (1%) | 11 (14%) |
| PPI | 19 (27%) | 55 (72%) |
| Allopurinol | 1 (1%) | 10 (13%) |
| Analgesic | 8 (11%) | 5 (7%) |
| Psychiatric | 7 (10%) | 10 (13%) |
| Other* | 22 (31%) | 69 (91%) |

Table 1- Participant characteristics – cohort for Analysis 1 (see Fig. 1)
| Characteristic | CNI-based triple therapy (n = 40) | mTORi-based triple therapy (n = 15) |
| --- | --- | --- |
| Age of transplant | 6.0 ± 5.1 (μ ± SD) | 14.8 ± 5.2 (μ ± SD) |
| Type of transplant | Deceased donor: 18 (45%)<br>Live unrelated: 13 (33%)<br>Live related: 9 (23%) | Deceased donor: 10 (67%)<br>Live unrelated: 4 (27%)<br>Live related: 1 (6%) |
| Graft number | First graft: 37 (91%)<br>Second graft: 3 (9%) | First graft: 11 (73%)<br>Second graft: 4 (27%) |
| Cause of ESKD |  |  |
| Glomerulonephritis | 17 (43%) | 9 (60%) |
| Other | 9 (22%) | 2 (13%) |
| Polycystic disease | 9 (22%) | 1 (7%) |
| Renovascular | 1 (3%) | 1 (7%) |
| Diabetic nephropathy | 2 (5%) | 0 (0%) |
| Unknown | 2 (5%) | 2 (13%) |
| Blood group | O+ 12 (30%)<br>O- 7 (18%)<br>A+ 14 (35%)<br>A- 4 (10%)<br>B+ 2 (5%)<br>AB+ 1 (3%) | O+ 5 (33%)<br>A+ 4 (27%)<br>A- 1 (7%)<br>B+ 3 (20%)<br>AB+ 2 (13%) |
| HLA mismatches |  |  |
| 0 | 2 (5%) | 0 (0%) |
| 1 | 5 (13%) | 2 (14%) |
| 2 | 5 (13%) | 1 (7%) |
| 3 | 8 (20%) | 2 (14%) |
| 4 | 10 (25%) | 2 (14%) |
| 5 | 7 (18%) | 5 (36%) |
| 6 | 3 (8%) | 2 (14%) |
| Induction therapy | Basiliximab: 19 (48%)<br>ATG: 18 (45%)<br>Eculizumab: 1 (3%)<br>Unknown: 2 (5%) | Basiliximab: 9 (64%)<br>ATG: 1 (7%)<br>Steroids alone: 2 (14%)<br>Unknown: 2 (14%) |
| Initial immunosuppression therapy | TAC/MMF/PRED: 39 (97%)<br>CyA/MMF/PRED: 1 (3%) | CNI/MMF/PRED: 7 (50%)<br>CyA/MMF/PRED: 4 (29%)<br>Belatacept/MMF/PRED: 1 (7%)<br>Unknown: 2 (14%) |
| Current immunosuppression therapy | TAC/MMF/PRED: 40 (100%) | mTORi/MMF/PRED: 15 (100%) |

Humoral immune response to vaccination was measured by total anti-Spike IgG titer, anti-receptor-binding domain (RBD) IgG titer, and live virus neutralization as the gold-standard correlate of protection from infection and severe disease (*24*). Cellular immunity was assessed by interferon (IFN)-γ ELISpot in response to stimulation with overlapping peptides spanning the SARS-CoV-2 Spike protein.

SARS-CoV-2-specific immunity was first compared between the complete KTR cohort and healthy controls (Fig. 2A, C, E). Consistent with previous reports, KTRs demonstrated significant impairment in anti-Spike IgG titer (median: AUC 1525 vs 0.8043, p<0.0001), seroconversion of anti-RBD IgG (100% vs 38.0%, p=0.0005), live virus neutralization (100% vs 18.3%, p<0.0001), and IFNγ-secreting Spike-specific T cell response (median: 430 vs 88 SFU/10^6^ cells, p<0.0001), irrespective of the vaccine type (ChAdOx or BNT162b2) (fig. S2).

**Fig. 2.**
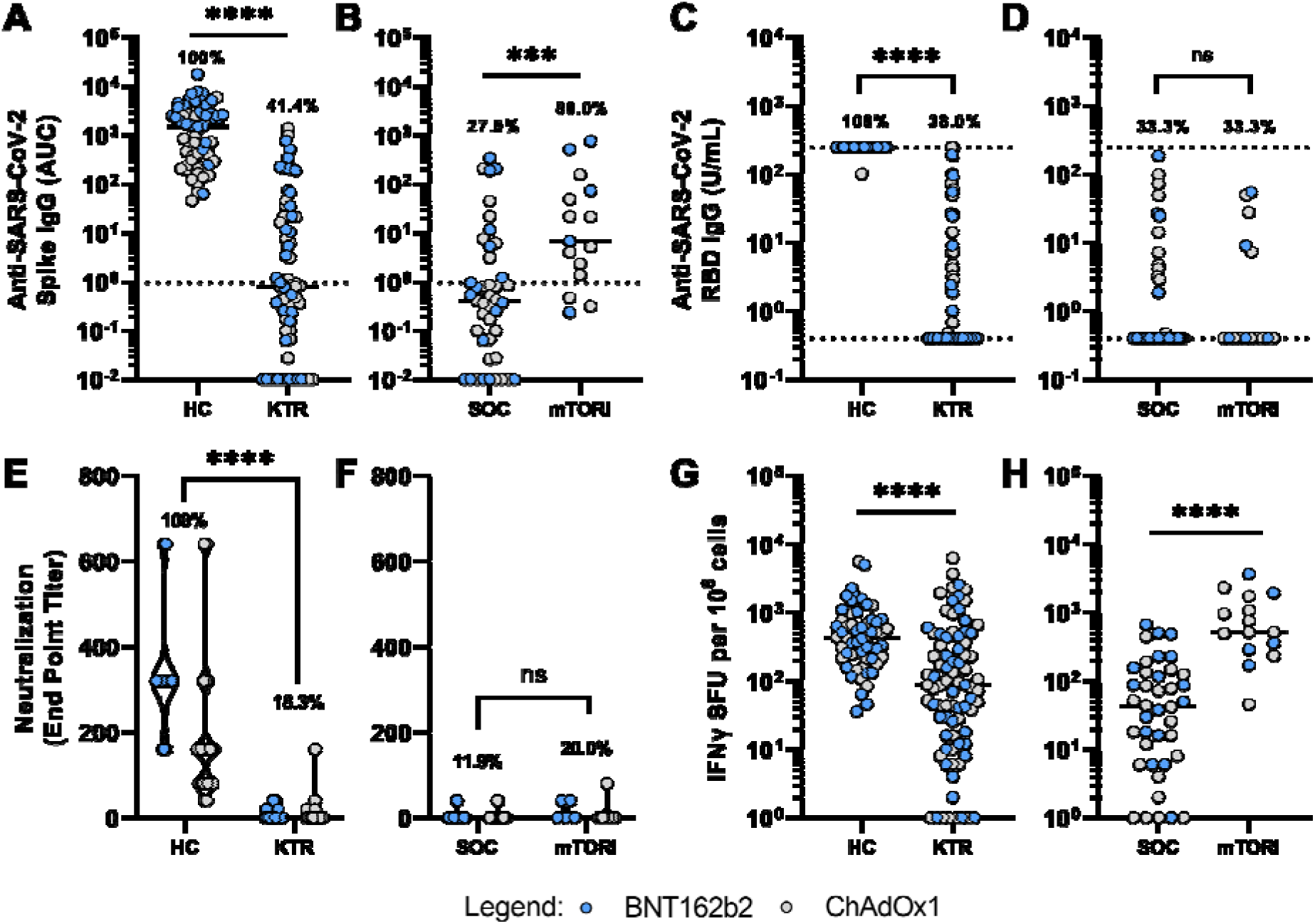
Immune response of kidney transplant recipients (KTRs) to a primary COVID-19 vaccination schedule. Responses of KTRs (n = 76) are compared against healthy controls (HC; n = 71), and between clinically comparable calcineurin inhibitor-based (‘SOC’; n = 43) and mTOR inhibitor-based (‘mTORi’; n = 15) three-drug immunosuppressive regimens. (**A-B**) End point titers are reported as area under the curve (AUC). Dashed lines represent detection threshold (mean AUC + 2 SD of negative controls). (**C-D**) Log-scale comparison of anti-RBD IgG. The detection range for the assay is 0.4 – 250 U/mL, as demarcated by dotted lines. Percentage of seropositive individuals (above 0.4 U/mL) is shown for each group. (**E-F**) Endpoint titers (minimum serum dilution) for 50% neutralization of live SARS-CoV-2 virus (ancestral strain) entry into target cells. (**G-H**) Log-scale comparison of Spike-specific IFNγ-secreting T cell responses measured as spot-forming units (SFU) by ELISpot. **Comparisons between groups by Mann-Whitney U test (flat bars) or Fisher’s exact test (crooked bars): ns, non-significant; *p<0.05; **p<0.01; ***p<0.001; ****p<0.0001.**

We next asked whether immunosuppression protocol has a significant effect on vaccine response. KTRs receiving standard-of-care (SOC) maintenance with a calcineurin inhibitor, anti-metabolite, and steroid were compared with those maintained on an alternative mTORi-based three-drug regimen (mTORi, anti-metabolite, steroid) (*15*), to assess which of these comparable protocols is preferable for an effective immune response to vaccination.

KTRs receiving the mTORi-based protocol (n = 15) demonstrated a median Spike-specific T cell response of 520 SFU/10^6^ cells, 12-fold higher than that of the SOC group (n = 43; p<0.0001), and similar to that of healthy controls (Fig. 2H). To confirm and further characterize the difference in T cell response between treatments, we evaluated the function and memory phenotype of Spike-specific CD4^+^ and CD8^+^ T cells in the mTORi group (n = 15), compared against a sub-cohort of KTRs on CNI-based SOC therapy (n = 15) and HCs (n = 15). Consistent with the ELISpot analysis, SOC immunosuppression was associated with impairment in the phenotype and function of Spike-specific effector memory T cells relative to healthy controls (fig. S3). Conversely, KTRs receiving the mTORi regimen formed T cell memory that was functionally and phenotypically similar to healthy individuals (fig. S3), and was superior to SOC in frequencies of all major CD4^+^ and CD8^+^ T cell memory populations, as well as greater frequencies of cytokine producing CD4^+^ and CD8^+^ T cells and stronger cytotoxic recall (GZMB^+^PRFN^+^) responses (fig. S3) (see Supplementary Materials).

Anti-Spike IgG titer was also significantly greater in mTORi-treated KTRs than SOC-treated (AUC 6.766 vs 0.4116, p=0.0011) (Fig. 2B). Despite this quantitative advantage, the two groups were equivalent in proportion of seroconverted patients with detectable anti-RBD IgG (33% vs 33%, p>0.99) (Fig. 2D), and in capacity of sera to neutralize live SARS-CoV-2 virus (ancestral strain A.2.2) (20.0% vs 11.9%, p=0.3910) (Fig. 2F).

The higher anti-Spike IgG titer for mTORi relative to SOC was surprising given the important role of mTORC1 signaling in the formation of T follicular helper (T_FH_) cells and germinal centers in mice (26). To investigate this, the antibody response in the sub-cohort was investigated in greater detail. In addition to the elevated anti-Spike IgG, we found a higher titer of anti-Spike IgM, and a trend towards greater anti-Spike IgA, for mTORi compared to SOC (fig. S4). Consistent with a role for mTORC1 in T_FH_ cell development, patients receiving mTORi demonstrated a reduced frequency of circulating (c)T_FH_ cells (defined as CD4^+^CXCR5^+^ T cells) versus SOC (fig. S4). However, a greater proportion of cT_FH_ cells in the mTORi group upregulated CD154 (CD40L) in response to stimulation with Spike peptides, suggesting that the reduced T_FH_ cell frequency may be partially offset by a greater proportion of antigen-specific T_FH_ cells capable of providing B cell help (fig. S3). No significant difference in live virus neutralization of Delta and Omicron BA.1 variants was observed between groups (fig. S4).

Taken together, usage of mTORi-based rather than SOC maintenance immunosuppression by KTRs was associated with superior induction of cellular and humoral immune responses to COVID-19 vaccination.

### KTRs receiving rapamycin have greater Spike-specific CD4^+^ and CD8^+^ memory T cell frequencies than healthy individuals

We next investigated whether mTORC1 inhibition promotes the formation of antigen-specific CD8^+^ memory T cells in humans, as previously described in animal models (*17, 18*). In support of this, KTRs on the mTORi-based triple therapy (mTORi, MMF, prednisolone) exhibited a trend towards a greater median IFNγ-secreting T cell response than healthy controls following primary vaccination (520 vs 430 SFU/10^6^ cells, p=0.1751; Fig. 2). We therefore extended our analysis to include all KTRs in our cohort who were receiving an mTORi as a component of two- or three-drug maintenance regimens, stratified by use of rapamycin (n = 14) or everolimus (n = 11) (Table 2) for comparison with healthy individuals.

**Table 2.** Participant characteristics – cohort for Analysis 2 (see Fig. 1)

| Characteristic | Healthy subgroup<br>(n = 15) | Rapamycin-containing regimen<br>(n = 14) | Everolimus-containing regimen<br>(n = 11) |
| --- | --- | --- | --- |
| **Age** | 54.1 ± 13.1 years | 61.5 ± 10.1 years | 66.9 ± 9.9 years |
| **Gender** | F: 12 (80%)<br>M: 3 (20%) | F: 5 (40%)<br>M: 9 (60%) | F: 2 (18%)<br>M: 9 (82%) |
| **Vaccine** | ChAdOx1-S: 6 (40%)<br>BNT162b2: 9 (60%) | ChAdOx1-S: 10 (71%)<br>BNT162b2: 4 (29%) | ChAdOx1-S: 6 (55%)<br>BNT162b2: 5 (45%) |
| **Graft number** | NA | First graft: 10 (71%)<br>Second graft: 4 (29%) | First graft: 9 (82%)<br>Second graft: 2 (18%) |
| **Cause of ESKD** | NA |  |  |
| Glomerulonephritis |  | 6 (40%) | 5 (45%) |
| Other |  | 1 (7%) | 1 (9%) |
| ADPKD |  | 2 (13%) | 3 (27%) |
| Renovascular |  | 1 (7%) | 1 (9%) |
| Unknown |  | 4 (26%) | 1 (9%) |
| **Current immunosuppression therapy** | NA | RAPA/MMF/PRED: 9 (64%)<br>RAPA/AZA/PRED: 1 (7%)<br>RAPA/MMF: 1 (7%)<br>RAPA/AZA: 1 (7%)<br>RAPA/PRED: 2 (14%) | EVERO/MMF/PRED: 6 (55%)<br>EVERO/AZA/PRED: 1 (9%)<br>EVERO/TAC/PRED: 3 (27%)<br>EVERO/MMF: 1 (9%) |
| **Comorbidities** |  |  |  |
| Hypertension | 6 (40%) | 13 (93%) | 10 (91%) |
| Diabetes mellitus | 1 (7%) | 3 (21%) | 2 (18%) |
| IHD | 0 (0%) | 2 (14%) | 0 (0%) |
| PVD | 0 (0%) | 0 (0%) | 0 (0%) |
| CerebroVD | 0 (0%) | 0 (0%) | 0 (0%) |
| Chronic lung disease | 1 (7%) | 0 (0%) | 0 (0%) |
| Dyslipidaemia | 1 (7%) | 12 (86%) | 8 (73%) |
| Cancer (inactive) | 2 (13%) | 3 (21%) | 4 (36%) |
| Kidney donor | 4 (27%) | NA | NA |
| Other | 6 (40%) | 12 (86%) | 8 (73%) |
| **Medications** |  |  |  |
| Antihypertensive | 4 (27%) | 12 (86%) | 10 (91%) |
| Lipid-lowering | 2 (13%) | 12 (86%) | 8 (73%) |
| Diabetic therapy | 1 (7%) | 1 (7%) | 2 (18%) |
| Antiplatelet | 0 (0%) | 2 (14%) | 3 (27%) |
| Anticoagulant | 1 (7%) | 5 (36%) | 4 (36%) |
| Anti-arrhythmic | 0 (0%) | 2 (14%) | 2 (18%) |
| PPI | 3 (20%) | 8 (57%) | 8 (73%) |
| Allopurinol | 1 (7%) | 0 (0%) | 3 (27%) |
| Analgesic | 1 (7%) | 1 (7%) | 0 (0%) |
| Psychiatric | 3 (20%) | 3 (21%) | 0 (0%) |
| Other* | 5 (33%) | 13 (93%) | 10 (91%) |

KTRs receiving rapamycin as a component of their immunosuppression regimen were found to have significantly greater frequencies of IFNγ-secreting, Spike-specific T cells than healthy individuals following primary COVID-19 vaccination (1020 vs 430 SFU/10^6^ cells; p=0.0395, Fig. 3A).

**Fig. 3.**
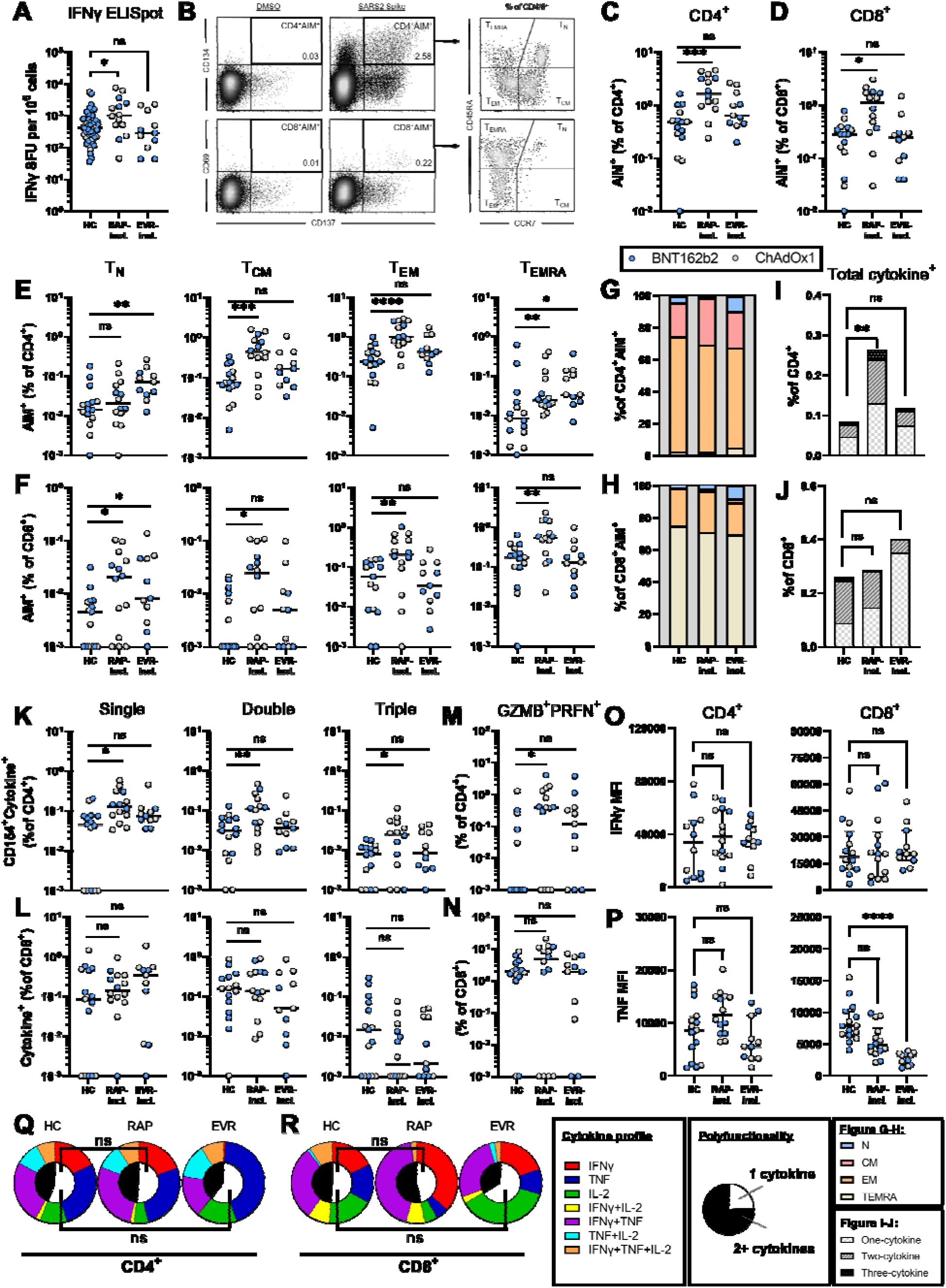
Phenotype and function of memory T cells following primary COVID-19 vaccination in healthy controls (HCs; n = 15) and kidney transplant recipients (KTRs) receiving rapamycin-inclusive (RAP-incl.; n = 14) and everolimus-inclusive (EVR-incl.; n = 11) immunosuppressive regimens. (**A**) Log-scale comparison of Spike-specific IFNγ-secreting T cell responses, measured as spot-forming units (SFU) by ELISpot. (**B**) T cells were gated as CD3^+^CD19 ^−^CD14^−^, and antigen-specific T cells defined by activation-induced marker (AIM) expression as CD4^+^CD134^+^CD137^+^ or CD8^+^CD69^+^CD137^+^. Within the AIM^+^ gates, four populations were defined by expression of CCR7 and CD45RA. (**C-D**) Percentage of AIM^+^ cells within the CD4^+^ (**C**) and CD8^+^ (**D**) T cell compartments. (**E-F**) Frequency of each AIM^+^ subpopulation as a percentage of the total CD4^+^ (**E**) or CD8^+^ (**F**) T cell populations. (**G-H**) Contribution of each subpopulation to the CD4^+^ (**G**) and CD8^+^ (**H**) AIM^+^ populations. Naïve: T_N_, CCR7^+^CD45RA^+^; Central Memory: T_CM_, CCR7^+^CD45RA^−^; Effector Memory: T_EM_, CCR7^−^CD45RA^−^; Terminally Differentiated Effector Memory: T_EMRA,_ CCR7^−^ CD45RA^+^. (**I-J**) Log-scale comparison of the percentage of CD4^+^ (**I**) and CD8^+^ (**J**) T cells positive for intracellular granzyme B (GZMB^+^) and perforin (PRFN^+^) following stimulation with Spike peptides. (**K-L**) Log-scale comparison of frequency of Spike-specific CD4^+^CD154^+^ (**K**) and CD8^+^ (**L**) T cells single, double, or triple positive for any combination of IL-2, TNF and IFNγ cytokines, as a percentage of CD4^+^ and CD8^+^, respectively. (**M-N**) Frequency of CD4^+^CD154^+^ (**M**) and CD8^+^ (**N**) T cells upregulating at-least one cytokine in response to stimulation, as a percentage of CD4^+^ and CD8^+^, respectively. (**O-P**) IFNγ (**O**) and TNF (**P**) mean fluorescence intensity (MFI) of Spike-specific CD4^+^CD154^+^ and CD8^+^ T cells. (**Q-R**) Median cytokine expression profile (outer pie chart) and proportion of cytokine positive cells that are polyfunctional (inner pie chart) of CD4^+^CD154^+^ (**Q**) and CD8^+^ (**R**) T cells. **Statistical analysis by Kruskal-Wallis with Dunn’s correction comparing each group to healthy controls: ns, non-significant; *p<0.05; **p<0.01; ***p<0.001; ****p<0.0001.**

No significant difference in T cell response was observed between the healthy and everolimus groups (Fig. 3A). It is unclear whether this reflects an intrinsic difference between the two mTOR inhibitors or the greater burden of co-immunosuppression in the everolimus group (Table 2).

To investigate the elevated T cell response associated with rapamycin, Spike-specific T cell frequencies were assessed independent of cytokine production by activation-induced marker (AIM) assay. For this analysis, isolated PBMCs were stimulated for 24h with a Spike-derived peptide array and Spike-specific T cells identified as CD4^+^CD134^+^CD137^+^ or CD8^+^CD69^+^CD137^+^ (*25, 26*) (fig. S5; Fig. 3B). KTRs receiving rapamycin exhibited a 3.3-fold higher median frequency of Spike-specific CD4^+^ T cells (1.60% vs 0.48%, p=0.0003), and a 3.1-fold higher frequency of CD8^+^ T cells (0.86% vs 0.28%, p=0.0017), than healthy individuals (Fig. 3C, D). This both reaffirms and extends our findings by ELISpot, demonstrating 1) that rapamycin promotes the formation antigen-specific memory T cells (rather than increasing IFNγ competency), and 2) affects both the CD4^+^ and CD8^+^ T cell compartments.

Memory T cell subpopulations within both the CD4^+^ and CD8^+^ compartments, defined by expression of CD45RA and CCR7, have overlapping and distinct roles in immunity, and in protection from SARS-CoV-2 in particular (*27–31*). We therefore assessed the frequencies of Spike-specific CD4^+^ and CD8^+^ T cells of a central memory (T_CM_; CCR7^+^CD45RA^-^), effector memory (T_EM_; CCR7^-^CD45RA^-^), terminally differentiated or terminal effector (T_EMRA_; CCR7^-^ CD45RA^+^), and naïve-like (T_N_; CCR7^+^CD45RA^+^) phenotypes (Fig. 3B, fig. S5). Rapamycin use did not appear to favor the formation of a particular memory phenotype (Fig. 3G, H), and KTRs on rapamycin exhibited greater frequencies of Spike-specific T_CM,_ T_EM_ and T_EMRA_ cells than healthy controls in both the CD4^+^ and CD8^+^ compartments (Fig. 3E, F). Rapamycin use was also associated with a significantly elevated frequency of Spike-specific CD8^+^CCR7^+^CD45RA^+^ ‘naïve’ T cells relative to healthy controls (Fig. 3E, F). While vaccination does not increase the frequency of antigen-specific naïve T cells, the CCR7^+^CD45RA^+^ phenotype also includes T stem cell memory (T_SCM_), a population which maintains the potential to differentiate into the full spectrum of T cell memory, and maintains long-term protection (*32*).

KTRs receiving everolimus as a component of their immunosuppression demonstrated a significant boost in the frequencies of Spike-specific CD4^+^ and CD8^+^ naïve-like, and CD4^+^ T_EMRA,_ cells (Fig 3E, F), but not in the total frequency of Spike-specific CD4^+^ and CD8^+^ T cells (Fig 3C, D).

These data suggest that rapamycin confers an improvement in the quantity of memory T cells that form in response to vaccination in humans. This effect was evident in both the CD4^+^ and CD8^+^ linages, and resulted in quantitative increases in the three major memory T cell compartments with defined roles in antiviral immunity. This trend was also evident at three months post-second vaccine dose in a subgroup of participants who had not received their third COVID-19 vaccine dose at follow-up (fig. S7).

### Spike-specific memory T cells associated with rapamycin use are highly functional

Previously, low-dose rapamycin treatment during the T cell contraction phase has been shown to improve the functionality of CD8^+^ memory T cells that arise in response to viral challenge - an effect that is uncoupled from the increase in memory CD8^+^ T cell frequency (*17*). Conversely, however, higher dose rapamycin, or genetic ablation of mTORC1, impairs the CD8 effector recall response (*17, 33*). We therefore assessed the functionality of Spike-specific memory T cells in the mTORi-treated KTRs compared to healthy individuals. Production of antiviral effector molecules IL-2, IFNγ, TNF, granzyme B (GZMB) and perforin (PRFN) in response to Spike peptide stimulation was assessed by intracellular cytokine staining (ICS) (Fig. 3I-R).

Functional Spike-specific T cells were enumerated as CD4^+^CD154^+^ or CD8^+^ cells producing one or more cytokines (IL-2, IFNγ, TNF) in response to stimulation (*34*). KTRs receiving rapamycin exhibited a greater frequency of single, double and triple cytokine-producing CD4^+^ T cells compared to healthy individuals, resulting in a marked increase in total frequency of cytokine-producing CD4^+^ T cells (Fig. 3I-L). This was consistent for all cytokines assessed individually (fig. S8). Although the frequency of AIM^+^ CD8^+^ T cells was greater in rapamycin treated KTRs than healthy individuals, the frequency of CD8^+^ T cells expressing each cytokine, and total cytokine positive cells, were similar between the groups (Fig. 3J, fig. S8B). KTRs in the everolimus group had comparable frequencies of functional Spike-specific CD4^+^ and CD8^+^ T cells to healthy individuals (Fig. 3B-L).

Simultaneous expression of multiple cytokines by memory T cells is associated with increased effector function and viral control (*35–40*). As a measure of functionality, the proportion of cytokine-producing cells that were polyfunctional, and the mean fluorescence intensity (MFI) of cytokines, was compared between healthy controls and KTRs on rapamycin or everolimus. Both mTORi groups demonstrated a similar proportion of cytokine-producing cells that were polyfunctional to healthy individuals (Figure 3Q, R) and, although there was a trend towards increased IFNγ and TNF MFI in CD4^+^ T cells in the rapamycin group, this did not reach significance (Fig. 3O, P). Both mTORi groups exhibited a trend towards lower TNF MFI in CD8^+^ T cells than healthy controls, reaching significance in the everolimus group (Fig. 3P).

As a measure of cytotoxic function, upregulation of intracellular granzyme B (GZMB) and perforin (PRFN) was measured upon stimulation with Spike peptides, to evaluate broader cytotoxic capacity of T cells in response to antigen encounter (*41*). KTRs on rapamycin had increased frequencies of CD4^+^ T cells that upregulated PRFN or PRFN^+^GZMB^+^ in resposne to antigen, while no difference in the frequencies of single or double GZMB/PRFN positive CD4^+^ and CD8^+^ T cells was observed for the everolimus group compared with healthy individuals (Fig. 3M, N; fig. S8).

Combinations of cytokine and effector molecules were further explored (fig. S9). IFNγ and TNF were the most commonly co-expressed molecules by CD4^+^ T cells, and granzyme B and perforin were most commonly co-expressed by CD8^+^ T cells (fig. S9). Penta-functional CD4^+^ T cells were detected in 40% (6/15) healthy individuals and 53% (8/15) KTRs on rapamycin (data not shown). Thus, following COVID-19 vaccination, the expanded pool of Spike-specific CD4^+^ and CD8^+^ memory T cells in KTRs on rapamycin is highly functional.

### Immunosuppression switch from MMF to rapamycin is safe and feasible but does not improve T cell response to COVID-19 booster vaccination in KTRs

The importance of T cell response in protection against SARS-CoV-2, and the clear benefit that rapamycin use had on the formation of functional T cell memory in our analysis, prompted the trial of immunosuppression modification with rapamycin as an intervention to enhance deficient T cell responses in KTRs to vaccination. An association between MMF use and severe impairment of seroconversion following COVID-19 vaccination in KTRs has been reported (*5*). We therefore hypothesized that switching MMF to rapamycin in KTRs on SOC therapy would both alleviate suppression of humoral immunity, and boost T cell responses to a third COVID-19 vaccine dose. The combination of CNI, rapamycin and steroid also minimizes the risk of rejection during the period of immunosuppression alteration relative to the combination of MMF, rapamycin and steroid, but does require a concurrent reduction in CNI dose to avoid nephrotoxicity (*42*). To test this, we initiated the RIVASTIM-Rapamycin trial (ACTRN12621001412820), a multi-center, randomized, open-label, controlled trial. We enrolled KTRs receiving triple therapy with tacrolimus, mycophenolate mofetil (MMF), and prednisolone, who exhibited a sub-optimal response (anti-RBD IgG<100 U/mL) following two vaccine doses (full criteria presented in S10). Participants were randomized to either cease MMF, start rapamycin 2 mg daily and reduce tacrolimus by 50% (rapamycin switch group), or to remain on existing immunosuppression (no switch control group) (see PRISMA diagram, fig. S11). Study protocol, methods and outcomes were pre-specified and were published elsewhere (*43*).

The trial was conducted from November 2021 – April 2022. Fifty-four patients were screened and all met criteria for randomization. A total of 28 patients were randomized to the rapamycin group and 26 to the control group. After attrition, 25 patients in the intervention group, and 23 patients in the control group, were included for outcome analyses. Patients were predominantly male (33/48, 69%), with mean age 59 ± 9.7 years, and had mostly received mRNA primary vaccination courses (28/48, 58%). Full baseline characteristics are shown in Table 3. In the rapamycin group, mean rapamycin trough concentrations were 6.3 ng/mL at time of vaccination, and 5.5 ng/mL at follow-up. Outcomes assessed included immune response to vaccine, including proportion achieving a protective viral neutralization titer threshold (primary outcome measure (*24, 43*)), anti-spike antibody titer, T cell response, and drug tolerability.

**Table 3.** Randomized, controlled trial baseline characteristics.

|  | Rapamycin Switch (n = 25) | No switch (n = 23) |
| --- | --- | --- |
| **Baseline Anti-RBD IgG** |  |  |
| Non-responder (< 0.4 units/mL) | 9 (36) | 7 (30) |
| Low-responder (≥ 0.4 units/mL) | 16 (64) | 16 (70) |
| **Age; mean (SD)** | 61 (8) | 56 (11) |
| **Sex** |  |  |
| Female | 8 (32) | 7 (30) |
| Male | 17 (68) | 16 (70) |
| **BMI at enrolment** |  |  |
| Normal (18.5-<25) | 5 (20) | 8 (35) |
| Overweight (25-<30) | 8 (32) | 6 (26) |
| Obese (30+) | 12 (48) | 9 (39) |
| **Self-reported ethnicity** |  |  |
| Aboriginal | 0 (0) | 1 (4) |
| Caucasian | 19 (76) | 19 (83) |
| Asian | 3 (12) | 2 (9) |
| Indian subcontinent | 1 (4) | 1 (4) |
| North African/Middle Eastern | 2 (8) | 0 (0) |
| **Time since transplant (years)** |  |  |
| 0-5 | 12 (48) | 10 (43) |
| 5-10 | 7 (28) | 5 (22) |
| >10 | 6 (24) | 8 (35) |
| **Initial COVID vaccine dose** |  |  |
| BNT162b2 (Pfizer) | 14 (56) | 13 (57) |
| ChAdOx1-S (Astra Zeneca) | 9 (36) | 9 (39) |
| mRNA 1273 (Moderna) | 1 (4) | 0 (0) |
| Heterologous vaccination | 1 (4) | 1 (4) |
| **Number of kidney transplants** |  |  |
| First graft | 20 (80) | 18 (78) |
| Second or greater graft | 5 (20) | 5 (22) |
| **Current Transplant type** |  |  |
| Deceased | 18 (72) | 16 (70) |
| Living | 7 (28) | 7 (30) |
| **Primary Renal Disease** |  |  |
| Diabetes mellitus | 3 (12) | 3 (13) |
| Polycystic kidney disease | 7 (28) | 4 (17) |
| Glomerulonephritis | 7 (28) | 8 (35) |
| Hypertension/renovascular disease | 1 (4) | 1 (4) |
| Other | 4 (16) | 4 (17) |
| Unknown | 3 (12) | 3 (13) |
| **Baseline eGFR; mean(sd)** | 55 (14) | 59 (20) |
| **Baseline urine A:Cr ratio** | 5 (8) | 14 (23) |
| **Baseline urine P:Cr ratio** | 12 (0) | 5 (0) |

The proportion of KTRs who exhibited protective neutralization titers was not different between groups (p = 0.85): rapamycin group (10/25, 40%) compared to the control group (9/21, 43%) (Fig. 4A). The unadjusted relative risk (RR) with rapamycin switch was 0.93 (95% CI 0.47 - 1.86, p = 0.84). This was also true of live virus neutralization for both ancestral (A.2.2) and Omicron BA.5 variant strains. No significant difference was observed in anti-RBD seroconversion in the rapamycin group compared to controls (12/25 vs 11/23, p = 0.82) (Fig. 4B), and this persisted after adjustment for baseline anti-RBD level (fig. S12, Table S3).

**Fig. 4.**
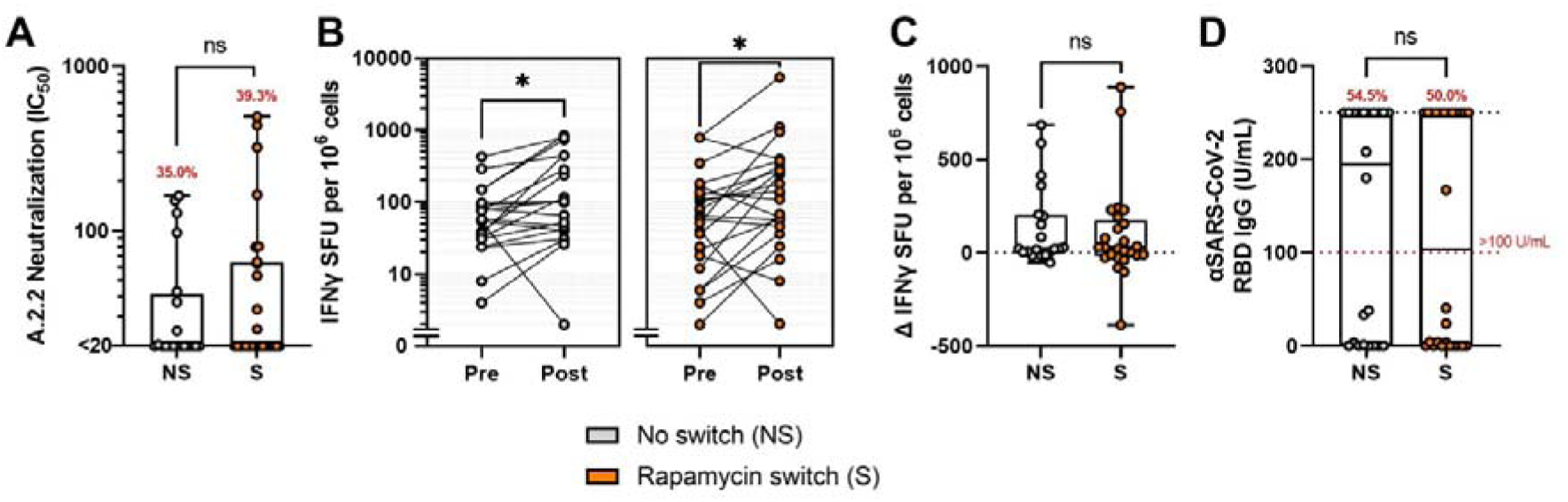
Immunological results of the RIVASTIM-Rapamycin trial. KTRs on SOC triple therapy who responded poorly (anti-RBD IgG<100 U/mL) to a primary two-dose vaccination schedule were randomized to the control ‘no switch’ or treatment ‘rapamycin switch’ trial arm four weeks prior to third dose of BNT162b2 COVID-19 vaccine. (**A**) Serum neutralization of live SARS-CoV-2 (ancestral A.2.2 strain) expressed as log IC_50._ (**B**) Spike-specific T cell responses pre- and post-booster vaccination, as measured by IFNγ ELISpot (SFU/10^6^ cells). There was a significant increase in response in both groups (p < 0.05). (**C**) Change in Spike-specific T cell response, as measured by change in IFNγ ELISpot (SFU/10^6^ cells). There was no significant difference in median change in T cell responses between ‘no switch’ and ‘rapamycin switch’ groups by quantile regression (p = 0.89). (**D**) Comparison of post-vaccination anti-RBD IgG titer (U/mL) between ‘no switch’ and ‘rapamycin switch’ cohorts. There was no significant difference in the proportion of patients reaching the pre-defined target threshold of 100 U/mL between groups (χ^2^ = 0.03, p = 0.87). **ns, non-significant.**

T cell responses as measured by IFNγ ELISpot increased following vaccination in both groups (p < 0.05, Fig 4C). However, the magnitude of change in T cell response did not differ between groups (p = 0.89, Fig 4D).

There was no significant difference in the frequency of any adverse events between the switch and control groups (27% vs 9%, p = 0.10), and there was no significant change in estimated GFR, and no episodes of transplant rejection over the course of the trial (Table S1-S2). Full details of secondary outcomes are available in the Supplementary Materials.

In summary, switching KTRs from MMF to rapamycin prior to a booster dose of COVID-19 vaccine was feasible and well-tolerated, however did not produce a positive effect on T cell response, as was apparent after primary COVID-19 vaccination for KTRs receiving rapamycin as a component of maintenance immunosuppression.

### Rapamycin enhances the frequency and function of antigen-specific memory T cells in mice

Immune response to booster vaccination is dominated by memory recall rather than *de novo* activation of naïve T cells (*44*). This, coupled with earlier reports that rapamycin influences memory formation by affecting the short-lived effector cell (SLEC) versus memory-precursor effector cell (MPEC) fate decision following activation of naïve T cells (*17, 19*), prompted us to investigate if rapamycin treatment differentially affects primary versus booster vaccine responses. To do so, we comprehensively evaluated the effect of rapamycin treatment before primary or booster COVID-19 vaccination in Balb/c mice (Fig. 1). Five groups of mice (n=7/group) were vaccinated with two doses of BNT162b2 vaccine (1 _μ_g/dose, IM), treated with rapamycin before primary dose, or before the third (booster) dose. To compare the effect of rapamycin on naïve versus recall response, mice were boosted with homologous (BNT162b2) or with plasmid DNA vaccine encoding Omicron SARS-CoV-2 variant RBD (pVAX-RBD-Omicron) (Fig. 1 and Fig. 5A).

**Fig. 5.**
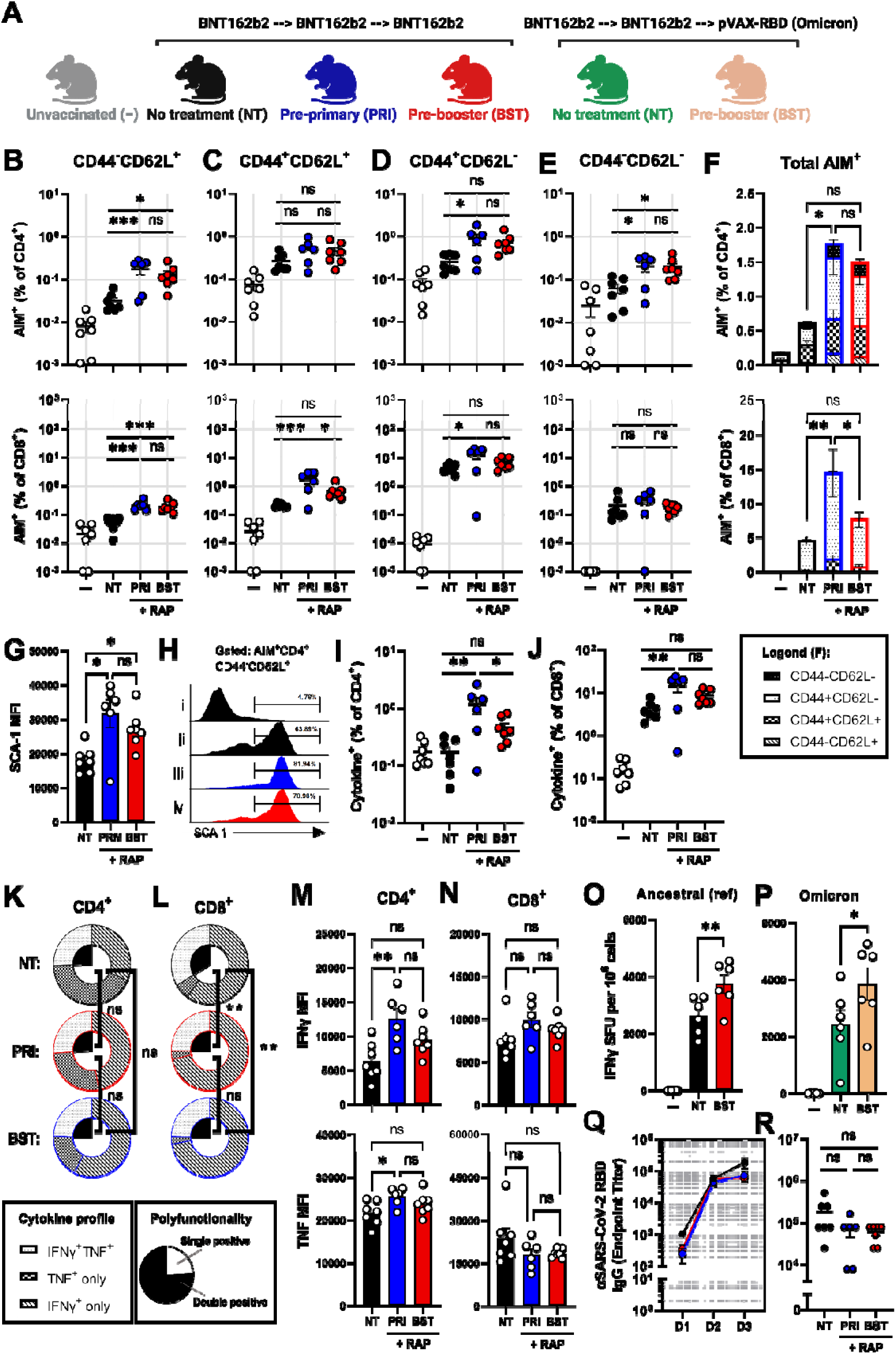
The effect of rapamycin on the immunogenicity of SARS-CoV-2 original and variant-strain vaccines is influenced by timing of treatment. (**A**) BALB/c mice received a primary vaccination schedule of BNT162b2 (Pfizer-BioNTech), followed by a booster dose of homologous (BNT162b2) or 50 ug of Omicron-specific (pVAX-RBD-Foldon[Omicron]) DNA vaccine. Mice were injected daily i.p. with 75μg/kg rapamycin beginning 5 days prior to the first dose (‘pre-primary’), or 5 days prior to booster vaccination (‘pre-booster’), or were not treated with rapamycin (‘no treatment’). Splenocytes were stimulated for 18 hours with peptides derived from the full-length Spike protein, and antigen-specific T cells defined by co-expression of the activation-induced markers (AIM) CD25 and CD69. (**B-F**) AIM^+^ T cells of (**B**) naïve (CD44^low^CD62L^+^), (**C**) central memory (CD44^high^CD62L^+^), (**D**) effector memory (CD44^high^CD62L^-^), and I double negative (CD44^low^CD62L^-^) phenotypes, and total AIM^+^ cells (**F**), were quantified as a percentage of CD4^+^ and CD8^+^ T cells. (**G-H**) Mean fluorescence intensity (**G**) and percentage (**H**) of stem cell antigen (SCA)-1 expression by AIM^-^ naïve-like (CD44^low^CD62L^+^) T cells (i) and AIM^+^ naïve-like (CD44^low^CD62L^+^) T cells from no treatment (ii), pre-primary (iii), and pre-booster (iv) rapamycin treated mice. I-N: Functional antigen-specific CD4^+^ and CD8^+^ T cells were assessed by expression of IFNγ and TNF following 16-hour stimulation with peptides derived from the full-length SARS-CoV-2 ancestral-strain Spike protein. (**I-J**) Percentage of CD4^+^ (**I**) and CD8^+^ (**J**) T cells expressing at-least one cytokine in response to stimulation. (**K-L**) Cytokine profile (outer pie chart) and polyfunctionality (inner pie chart) of Spike-specific CD4^+^ (**K**) and CD8^+^ (**L**) T cells. (**M-N**) Mean fluorescence intensity of cytokine expression by IFNγ^+^ and TNF^+^ T cells in the CD4^+^ (**M**) and CD8^+^ (**N**) compartments. (**O-P**) The effect of rapamycin treatment beginning pre-booster dose on wildtype (**O**) and Omicron-specific (**P**) T cell responses to the BNT162b2 and pVAX-RBD-Foldon[Omicron] vaccines, respectively. (**Q**) Serum titers of anti-RBD IgG following the first (D1), second (D2) and third (D3) vaccine doses. (**R**) Effect of rapamycin treatment from pre-primary or pre-booster time points on the final (D3) anti-RBD IgG titer. **Multiple comparisons by one-way ANOVA with Holms’ correction: ns, non-significant; *p<0.05; **p<0.01; ***p<0.001; ****p<0.0001.**

Supporting the findings in KTRs, administration of rapamycin from baseline resulted in significant improvement in the frequency of functional Spike-specific T cell memory populations following three doses of BNT162b2 vaccine (Fig. 5). Similar to patients, rapamycin treated mice had significantly higher frequencies of Spike-specific CD4^+^ and CD8^+^ T cells by AIM assay (Fig. 5F; fig. S13). This increase was evident for CD4^+^ T_EM_ and CD44^low^CD62L^-^ populations, and for CD8^+^ T_EM_ and T_CM_ populations (Fig. 5C, D). Increased frequencies of AIM^+^ cells of a naïve-like (CD44^low^CD62L^+^) phenotype were observed in both the CD4^+^ and CD8^+^ T cell compartments, mirroring the increased frequencies of AIM^+^ naïve-like (CD45RA^+^CCR7^+^) CD8^+^ population in KTRs on rapamycin (Fig. 5B; Fig. 3E, F). In contrast to the bulk (AIM^-^) naïve-like T cells, this antigen-specific population expressed high levels of SCA-1, suggestive of a T_SCM_ identity (Fig. 5G, H). Stem-like memory T cells have also been reported to express additional memory markers such as CXCR3; AIM^+^CD44^low^CD62L^+^SCA-1^+^ cells in our study were negative for CXCR3, and thus embody a partial T_SCM_ phenotype (fig. S14). mTORC1 inhibition has been reported to increase T_SCM_ formation *in vitro* (*45*), and the percentage of AIM^+^CD44^low^CD62L^+^ cells positive for SCA-1, and the MFI of SCA-1 staining, was significantly higher in rapamycin treated mice (Fig. 5G, H).

Rapamycin treated mice exhibited increased frequencies of CD4^+^ and CD8^+^ cytokine-producing T cells (Fig. 5I, J), as well as increased frequencies of multi-functional (TNF^+^IFNγ^+^) CD4^+^ and CD8^+^ T cells (Fig. 5L, K). Similar to KTRs (Fig. 4R), rapamycin favored the formation of IFNγ single-positive T cells (Fig. 5L, K). This resulted in a smaller proportion of cytokine-producing CD8^+^ T cells being multi-functional (TNF^+^IFNγ^+^) despite the increased frequency of these cells (Fig. 5L, K). In support of an additional beneficial effect of mTORC1 inhibition on memory T cell functionality, rapamycin treatment increased the average MFI of both TNF and IFNγ expression by cytokine positive CD4^+^ T cells (Fig. 5M). Of note, while it did not reach significance, a small reduction in the MFI of TNF expression by TNF^+^CD8^+^ T cells was observed that mirrored a similar finding in the KTRs receiving mTORi compared with healthy individuals (Fig. 5N; Fig. 3P).

Considerable variation in T cell response was observed in the rapamycin-treated group, particularly in the CD8 compartment (Fig. 5). Rapamycin concentrations were measured in serum at the time of T cell analysis and were found to inversely correlate with CD8^+^ T cell response by AIM (r = −0.8696, p = 0.0244) and cytokine production (r = −0.9346, p = 0.0063) (fig. S14). This negative association is consistent with reports that high dose rapamycin or genetic ablation of mTORC1 impair memory T cell function at recall (*17, 33*).

To evaluate the effect of rapamycin treatment on the humoral immune response in mice, we measured anti-RBD IgM and IgG titers. Following the first vaccine dose, mice receiving rapamycin demonstrated significantly higher titers of anti-RBD IgM, and lower titers of anti-RBD IgG (fig. S14), in line with previously reported effects of rapamycin (*46, 47*). These effects diminished considerably, however, with repeat vaccination, such that no significant differences were observed between groups following the second and third doses, for either antibody isotype (Fig. 5; fig. S14).

### T cell responses to SARS-CoV-2 original and variant-strain vaccines are influenced by timing of rapamycin treatment in mice

To understand the lack of effect of rapamycin in our randomized, controlled trial, a comparator group of mice were started on rapamycin after two doses of vaccine (prior to administration of the third dose) to evaluate the potential of rapamycin to improve the response to booster vaccination (Fig. 1). Consistent with the conflicting outcome of our observational and interventional studies in KTRs, beginning rapamycin treatment prior to booster vaccination produced a modest, but non-statistically significant, increase in the frequency of Spike-specific CD4^+^ and CD8^+^ T cells upregulating activation markers and antiviral cytokines in response to stimulation (Fig. 5F, I, J). Significant increases were observed in Spike-specific naïve-like T cells in the both CD4^+^ and CD8^+^ compartments, as well as elevated SCA-1 expression in this population, indicating a partial effect relative to rapamycin treatment from baseline (Fig. 5B, G).

As rapamycin is thought to influence the fate of activated naïve T cells (*17*), a predominant process of primary rather than recall immunity (*48*), we next evaluated if the effect of rapamycin is more pronounced for novel antigens. The implication of this, given that the majority of the population has been exposed to SARS-CoV-2 via vaccination or infection, would be that rapamycin treatment could preferentially enhance the response to novel epitopes in variant vaccines. We therefore assessed the response to an Omicron variant RBD plasmid DNA vaccine (pVAX-RBD-Foldon[Omicron]) in mice that had received two doses of BNT162b2 vaccine (Fig. 1). The effect of rapamycin on the naïve T cell response (response to Omicron-specific epitopes in Omicron-boosted mice) and the recall T cell response (response to equivalent ancestral epitopes in homologous boosted mice) were compared. Responses were assessed by IFNγ ELISpot to maximize sensitivity for detection of novel epitope-specific T cells.

Beginning rapamycin treatment prior to boosting with the Omicron RBD vaccine produced a significant 1.58-fold increase in IFNγ-secreting Omicron-specific T cells relative to mice that did not receive rapamycin (Fig. 5P). While this suggests that rapamycin administration at the time of variant booster may benefit the T cell response, a similar (p = 0.4332) 1.43-fold increase in the T cell response to ancestral Spike epitopes was produced by rapamycin treatment of mice peri-booster vaccination with the homologous BNT162b2 vaccine (Fig. 5O). Thus, contrary to the hypothesis that mTORC1 inhibition preferentially enhances T cell responses to novel epitopes, rapamycin had a similar, modest effect on the T cell response to both homologous and variant vaccine boosters.

Collectively, while rapamycin commenced prior to booster vaccination enhanced the T cell response and promoted the formation of T_SCM_-like cells, these effects on cellular immunity were significantly greater when rapamycin was commenced prior to initial vaccination.

## DISCUSSION

In the present study, KTRs receiving rapamycin at the time of primary COVID-19 vaccination formed greater frequencies of SARS-CoV-2 Spike-specific T cells than healthy individuals. This effect of mTORC1 inhibition on the T cell response to vaccination, which has been reported previously in preclinical models (*17, 33, 49-51*) and is described here for the first time in humans, was characterized by increased antigen-specific CD4^+^ and CD8^+^ T cells of the three major memory subsets (T_CM_, T_EM,_ and T_EMRA_), as well as elevated frequencies of polyfunctional T cells. These findings were similar to the effect of rapamycin treatment on COVID-19 vaccinated mice, where rapamycin augmented the CD4^+^ and CD8^+^ T cell memory responses to primary vaccination, and was associated with the expansion of a Spike-specific T_SCM_-like population in mice and humans. Comparison of KTRs on an mTORi-based versus SOC three-drug maintenance regimen found a 12-fold greater IFNγ ELISpot response to primary vaccination, demonstrating the potential benefit of mTORC1 modulation as a strategy to improve T cell response to vaccination of KTRs. In contrast to the SOC group, antigen-specific T cells in these patients exhibited a healthy distribution between memory subsets, and the frequency of CD4^+^ and CD8^+^ T cells producing IFNγ, TNF, IL-2, granzyme B and perforin in response to Spike peptides was also higher in the mTORi-based therapy group.

Inadequate response to vaccination is a major problem facing immunocompromised KTRs, underscored by the 24% mortality rate early in the COVID-19 pandemic (*1*), and by the limited protection afforded this group by standard COVID-19 vaccination strategies (*3*). A number of strategies have been employed to try to improve protective immunity in KTRs, which predominantly focus on enhancing or maintaining humoral immunity to SARS-CoV-2, including multiple vaccine boosters (*5, 6, 10, 11, 52-54*), as well as ring vaccination (*13, 55*), convalescent plasma and immunoglobulin (*56, 57*), and prophylactic monoclonal antibody therapy (*58, 59*). While repetitive booster vaccinations have shown benefit (*4, 7, 11, 54*), there are limitations to this strategy (*60–62*), and a subgroup of KTRs, particularly those receiving higher doses of the anti-metabolite mycophenolate (mycophenolate mofetil, MMF or mycophenolic acid, MPA), remain seronegative (*5*).

Less attention has been given to improving the T cell response, despite severe impairment in cellular immune responses to COVID-19 vaccination associated with standard immunosuppression in KTRs (*5, 13*). Improving the T cell response is a long-standing goal of vaccine development, as effective T cell immunity can confer protection against severe disease from viral infection, and cross-protection against antibody-escape variants (*2, 34, 63*), which is particularly important in immunocompromised groups, such as kidney transplant recipients, that lack effective neutralizing antibody responses (*64*). The importance of robust vaccine induced cellular immunity to COVID-19 was recently highlighted in an open letter to the U.S. Food and Drug Administration (FDA) stressing the need for standardized research into T cell immunity (*65*).

In addition to the improved T cell responses, patients receiving the mTORi-based three-drug regimen at the time of vaccination generated significantly higher titers of anti-Spike IgM and IgG than SOC treated KTRs. While the elevated titers of anti-Spike IgM observed in the mTORi group are consistent with previous observations in mice treated with rapamycin peri-vaccination (*46, 47*), the higher titers of anti-Spike IgG were unexpected, as mTORC1 signaling is known to be important for the germinal centre response and, by extension, for T_FH_ differentiation and class-switching (*46*). A reduction in cT_FH_ cell frequency in KTRs on the mTORi-based regimen was observed, however was offset by an increased frequency of Spike-specific cT_FH_ cells, and no reduction in anti-RBD IgG levels was found in vaccinated mice treated with rapamycin. Thus, as CNIs also impaired the antibody response (*66, 67*), the relative increase in anti-Spike IgG in the mTORi group may simply reflect greater suppression of the anti-Spike IgG response associated with CNI use rather than mTORi use. Alternatively it is possible that as individuals in this study were older, any negative impact of mTORC1 inhibition on the humoral immune response might be offset by reversal of immune senescence, as has been previously demonstrated in randomized controlled trials using mTORC1 inhibitors (*68, 69*). In a recent observational study, a similarly elevated IgG response to COVID-19 vaccination was reported for KTRs receiving CNI, mTORi, and prednisolone, compared with those receiving standard-of-care (*70*). Given the known effects of MMF on antibody response to COVID-19 vaccination, MMF rather than rapamycin use likely accounts for the observed difference (*5, 71–73*).

The clear benefit of rapamycin treatment on the T cell response in COVID-19 vaccination prompted investigation of immunosuppression modification with rapamycin as a strategy to improve the response of KTRs to COVID-19 booster vaccination. In the randomized, controlled trial, performed during the pandemic, MMF was withdrawn from KTRs on SOC immunosuppression who responded poorly to the first two vaccine doses, and was replaced with rapamycin prior to booster dose vaccination. As a pilot trial, participant numbers were small and inadequately powered to detect a difference in the primary outcome of neutralization. Several trials have been conducted recently that withdraw MMF or MPA prior to booster vaccination with the aim of improving the antibody response in non-responsive KTRs (*74–77*), however no clear benefit to booster vaccine response has been demonstrated.

The trial was, however, sufficiently powered to detect a meaningful difference in T cell ELISpot between the groups based on our observational findings. Despite this, no significant difference in the magnitude of response from pre- to post-vaccination, nor in final ELISpot counts, was observed. There were several differences in the circumstances of the observational and interventional trials that might account for the different outcomes. The first is the effect on primary versus booster vaccination.

As the majority of patients had received booster vaccinations or had had COVID-19, the vaccine response was studied in mice to evaluate whether rapamycin confers a significant effect on T cell response at the time of booster vaccination. While we found evidence of an improvement in T cell response to booster vaccination with rapamycin treatment, the effect was modest compared to the effect when administered from baseline.

Although the effect of rapamycin treatment on the T cell response to booster vaccination was limited, it was hypothesized that a greater effect would be observed to the novel epitopes in an Omicron-specific booster vaccine, as mTORC1 is thought to confer its effect by promoting memory-precursor effector cell (MPEC) formation from activated naïve T cells (*78*). To the contrary, naïve and recall responses were enhanced to a similar extent by rapamycin and, as such, a cumulative effect of rapamycin over multiple vaccine doses may instead account for the differences that were observed in response with timing of treatment. Rapamycin treatment prior to a booster vaccination did, however, result in a significant increase in Spike-specific T_SCM_-like T cells in mice.

Another factor that may have contributed to the lack of effect in the RCT was the approach to immunosuppression modification. The observational cohort consisted predominantly of KTRs on mTORi, MMF and corticosteroid, however we elected to switch participants of the RCT to a tacrolimus, mTORi, corticosteroid regimen, in order to minimize risk of transplant rejection, and to relieve suppression of the antibody response by MMF. Mutual antagonism between tacrolimus and rapamycin has been reported due to shared intracellular binding to FK-binding proteins, however these drugs are synergistic at the concentrations used *in vivo* (*79*). Tacrolimus has also been reported to promote the formation of short-lived effector T cells (SLECs) in mice, which occurred at the expense of the formation of MPECs (*80*). As rapamycin is thought to also act on this MPEC vs SLEC junction, but in the opposite direction, a mechanism exists by which tacrolimus might interfere with the effect of rapamycin on T cell memory formation, and have contributed to the lack of effect in our RCT. Despite the theoretical potential for this drug-drug interaction, the combination of everolimus and tacrolimus has been reported to associate with a lower incidence of viral infection than tacrolimus and mycophenolate in human KTRs (*16*).

This study was limited in its reliance on comparison with healthy individuals. The conclusion that rapamycin use confers an improvement in the formation of functional T cell memory is predicated on the assumption that the other immunosuppressants received by the patients in the rapamycin-inclusive group, and their status as kidney transplant recipients more broadly, has a neutral or negative influence on the T cell response relative to the healthy control group. While we believe this assumption is reasonable, it precludes accurate assessment of the magnitude of the observed effect in humans. Due to restraints on participant recruitment during the height of the COVID-19 pandemic, recruitment targets for the RIVASTIM-Rapamycin trial were not met, and as such, conclusions on the efficacy of MMF to rapamycin switch on the primary outcome of serological neutralization could not be drawn.

In conclusion, modulation of mTORC1 is a valid strategy to improve the T cell memory response to vaccination in humans. Low-dose rapamycin at the time of primary COVID-19 vaccination augmented the formation of key CD4^+^ and CD8^+^ T cell memory populations, with the capacity for a highly functional antiviral recall response. These benefits of mTORC1 inhibition were observed in KTRs receiving an mTORi-based regimen at the time of primary COVID-19 vaccination, who did not exhibit the severe impairment in T cell response that was associated with standard immunosuppression. In the absence of an effective neutralizing antibody response, a functional T cell response in these patients may provide protection from severe disease and cross-protection against variant strains (*34, 64, 81*). While further studies are needed to define the optimal context for the application of mTORi to improve vaccine responses in KTRs, low-dose rapamycin treatment peri-vaccination was feasible and well-tolerated in this study. In conjunction with recent trials of mTORC1 inhibitors in the elderly, there is now clear evidence of the benefit and feasibility of short-term mTORi treatment as a widely applicable T cell adjuvanting therapy.

## MATERIALS AND METHODS

### Study design: observational

Participants were prospectively recruited for the REVAX trial (ACTRN12621000532808), for which a partial cohort was previously published (*13*). This study was conducted on a single center population of KTRs and their healthy cohabitants commencing in February 2021. Participants were identified by clinical nephrologists and screened. Inclusion criteria were kidney-only transplant over 18 years old with an available cohabitant without kidney disease. Exclusion criteria were patients with past COVID-19 infection, those who had already received a COVID-19 vaccine prior to enrolment, those who could not provide informed consent, or those who did not have a non-immunosuppressed and unvaccinated cohabitant. Demographics including gender, age, cause of kidney disease, and graft details were collected and presented as percentages for ordinal variables, and median ± range for continuous variables.

For AIM and ICS analyses, 15 HC and 15 SOC KTRs were sampled from the complete cohort. This was done using a random number generator (Random.org), which produces true randomness based on atmospheric noise. No experiments were blinded in this study.

### Study design: interventional

The RIVASTIM trial was a prospective, multicenter, randomized, controlled trial performed at two academic transplant centers in Australia. Details regarding the trial design and rationale were pre-specified and published previously (*43*).

#### Study population

From 8 November 2021 until 15 February 2022, patients were recruited and screened for eligibility. All eligible patients were required to be kidney transplant recipients 18 – 75 years of age on standard immunosuppressive therapy with tacrolimus, mycophenolate, and prednisolone. Patients required good graft function with estimated GFR >25 mL/min/1.73m2, spot urinary albumin:creatinine ratio <100 mg/µmol, and must have received 2 doses of a COVID-19 vaccine with suboptimal immune response as defined by anti-RBD IgG <100 U/mL. Full inclusion and exclusion criteria are outlined in fig. S10.

#### Trial procedure

Eligible participants were randomized 1:1 stratified by study site and initial immune response (non-response <0.4 U/mL vs. low-responder 0.4-99 U/mL) with randomly permuted blocks of 2, 4 and 6. Patients in the control group continued their usual immunosuppression regimen of tacrolimus, mycophenolate, and prednisolone. Patients in the intervention group ceased mycophenolate and commenced rapamycin (sirolimus) 2 mg daily, titrated weekly to trough levels 6 ng/mL. Tacrolimus dose was also adjusted to achieve trough levels 3-6 ng/mL. After a 4-week lead in period, participants received a 3rd dose of mRNA COVID-19 vaccine, with subsequent antibody response measured at 4-6 weeks post-vaccination.

#### Trial outcomes

The primary trial outcome was the proportion of participants in each trial arm who developed protective serological neutralization of live SARS-CoV-2 virus (defined as 20.2% of the mean neutralization level of a standardized cohort of COVID-19 convalescent individuals). Secondary outcomes were measures of humoral immunity based on anti-RBD IgG, and measures of cellular immunity based on ELISpot counts.

#### Recruitment

The study aimed to enroll 120 patients across both sites, with 60 assigned to each group. This would provide 80% power (alpha 0.05) to detect an absolute difference of 25% in the proportion of patients achieving the neutralization threshold, allowing for a 10% drop-out rate. Recruitment was closed early due to the rapid reduction in eligible trial participants with the ongoing rollout of booster COVID-19 vaccines, combined with the increasing community prevalence of COVID-19 that made further delays in booster doses undesirable.

### SARS-CoV-2 Spike protein production and ELISA

Prefusion SARS-CoV-2 Spike ectodomain (isolate WHU1, residues 1-1208) with HexaPro mutations (kindly provided by Dr Adam Wheatley)(*82*) and SARS-CoV-2 RBD with C-terminal His-tag (residues 319-541; kindly provided by Prof Florian Krammer) were used in ELISA as we previously described (*34, 83*). Patient sera were serially diluted and end point titers of SARS-CoV-2 Spike and RBD were calculated and expressed as area under the curve (AUC) using the mean optical density (OD) reading of the first dilution from healthy normal human sera as the baseline cut-off for seropositivity. AUC calculations were performed using Prism GraphPad.

‘Elecsys Anti-SARS-CoV-2 S’ and ‘Elecsys antiSARS-CoV-2’ assays on the Cobas system (Roche), were used to quantify RBD- and nucleocapsid IgG as per manufacturer’s instructions. The quantitation range for detection of anti-RBD IgG in this assay is 0.4-250 U/mL. One U/mL is comparable to 1 BAU/mL.

### IFNγ ELISpot

Thawed PBMCs were stimulated for 18h with 4 pools of overlapping peptides spanning the entire length of the spike glycoprotein of the USA-WA1/2020 strain. Individual peptides are 17- or 13-mers, with 10 amino acid overlaps, obtained through BEI Resources, NIAID, NIH (*Peptide Array, SARS-Related Coronavirus 2 Spike (S) Glycoprotein, NR-52402*).

Briefly, multiscreen-IP HTS plates (Merck Millipore) were coated with anti-human IFN-γ (clone 2G1, Thermo Fisher), and secreted IFN-γ was detected with anti-human IFN-γ biotin (Clone B133.5; ThermoFisher) followed by streptavidin–HRP (BD Biosciences) and AEC substrate (BD Biosciences). Developed spots were counted automatically by use of an ELISpot reader (Cellular Technology Ltd., Bonn, Germany), and the number of spots for unstimulated splenocytes (<50) was subtracted from the number of spots for the peptide pool–stimulated splenocytes to generate the number of specific spot-forming units per 10^6^ cells. Data are presented as mean ± SEM.

### SARS-CoV-2 live-virus neutralization assay

Rapid high-content neutralization assay with HEK-ACE2/TMPRSS cells, HAT-24, cells was done as previously described (*84, 85*). HAT-24 cells were seeded in 384-well plates at 16 × 10^3^ cells/well in the presence of the live cell nuclear stain Hoechst-33342 dye (NucBlue, Invitrogen) at a concentration of 5% v/v. Two-fold dilutions of patient serum samples were mixed with an equal volume of SARS-CoV-2 virus solution standardized at 2xVE_50_ and incubated at 37°C for 1 h before adding 40 μL, in duplicate, to the cells. Viral variants used included key variants of concern, Delta (B.1.617.2), Omicron (B.1.1.529) and Omicron sub-variant BA.5, as well as ‘ancestral’ virus (A.2.2) from clade A and presenting no amino acid mutations in Spike (similar to Wuhan ancestral variant). Plates were incubated for 24 h post infection and entire wells were imaged by high-content fluorescence microscopy, cell counts obtained with automated image analysis software, and the percentage of virus neutralization was calculated with the formula: %N = (D-(1-Q)) × 100/D, as previously described (*84, 85*). Sigmoidal dose-response curves and IC_50_ values (reciprocal dilution at which 50% neutralization is achieved) were obtained with GraphPad Prism software.

### SARS-CoV-2 Spike peptide pools for AIM and ICS

The SARS-CoV-2 peptide pool used for AIM and ICS analyses was kindly provided by Prof Alessandro Sette (La Jolla Institute of Immunology, CA, USA) (*86*). 15-mer peptides overlapping by 10 amino acids and covering the entire Spike protein sequence were used (total of 253 peptides). All peptides were synthesized and resuspended in DMSO at 1 mg/ml. Thawed PBMCs were rested for 2 h and stimulated with 1 μg/mL of SARS-CoV-2 Spike megapool. PHA 10 μg/mL (Sigma Aldrich) was included as a positive control. An equimolar amount of dimethyl sulfoxide (DMSO, vehicle) was used as a negative control. PBMCs were stimulated for 24 h at 37°C, 5% CO_2_, washed and stained with Zombie Green Fixable Live/Dead Stain (L/D, Biolegend) for 20 min, RT, in the dark. Cells were then washed and stained with (CD3 BUV737, CD4 BUV496, CD8 BUV395, CD14 FITC, CD19 FITC, CD45RA BV650, CCR7 (CD197) APC, CD69 PE, CD134 (OX40) PE-Cy7, CD137 (41-BB) BV421) for 20 min, at room temperature in the dark. Fluorescence minus one (FMO) control for antigens: CD45RA, CCR7, CD134, CD69 and CD137 were added to PHA stimulated cells. PBMCs were washed and FACS Fix (0.4%PFA, 20 g/L Glucose, Sodium Azide 0.02% in PBS) was added for 20 min at room temperature, in the dark. Fixed cells were washed, resuspended in FACS wash buffer and data was acquired on BD FACS Symphony. Data analysis was performed using FCS Express ™ (DeNovo Software, Pasadena, CA, USA). All percentages of activated cells were calculated subtracting unspecific DMSO background for each cell phenotype and individual patient.

### Intracellular cytokine staining and Spike-specific T follicular helper cell quantification

As described previously (*87*), PBMCs were thawed and prepared for cell culture as described for the AIM assay. Cells were pre-treated with 0.555 μg/mL of anti-CD40 blocking antibody (HB14, Miltenyi Biotec) for 15 min. Spike peptide pool was then added to a final concentration of 1 μg/mL (making anti-CD40 concentration 0.5 μg/mL for the remainder of the stimulation period). After a 20 h incubation, 2 μM GolgiStop™ (containing monensin, BD, 554724) and 1 μg/mL GolgiPlug™ (containing Brefeldin A, BD, 555029) were added to the cells and incubated for an additional 4 h. Cells were then co-incubated with Fixable Viability stain 780 (BD) and Fc Block (BD) for 20 min, RT, in the dark, washed with FACS buffer solution and stained with surface stain mix (CD3 BUV737, CD4 BUV496, CD8 BUV395, CXCR5 BUV563, CD14 APC-Cy7, CD20 APC-Cy7) for 20 min, RT, in the dark. Cells were washed with PBS and subsequently fixed and permeabilized with Cytofix/Cytoperm™ (BD, 51-2090KZ) for 20 min, RT, in the dark. Cells were then washed with Perm/Wash™ (BD, 51-2091KZ) and stained with ICS stain Mix (CD154 PE, IFNγ PE-Cy7, TNFα APC, PRF1 FITC, IL-2 BV711, GZMB BV421) for 20 min, RT, in the dark. Cells were then washed twice with Perm/Wash™ and once with PBS. Finally, cells were resuspended in PBS and kept at 4°C until data was acquired on BD FACS Symphony. Data analysis was performed using FCS Express ™ (DeNovo Software, Pasadena, CA, USA).

### Animals

All mouse work was conducted in accordance with the *Australian Code for the Care and use of Animals for Scientific Purposes* as defined by the National Health and Medical Research Council of Australia, and studies were approved by the University of Adelaide Animal Ethics Committee. Forty-two female, 6-8-week-old, Balb/c mice were purchased from the University of Monash and randomly allocated to groups (n=7 per group). Balb/c mice were vaccinated intramuscularly with two doses of 1ug of BNT162b2 (Pfizer) vaccine at three-week intervals. Six weeks after the second dose, mice received the third dose of BNT162b2 vaccine (1ug) or 50 ug of pVax-RBD (omicron) DNA vaccine intradermally into the ear pinnae.

Rapamune oral solution (sirolimus, 1 mg/mL) was administered with daily intraperitoneal injection of 0.075mg/kg rapamycin in 100 uL (*17*). The dosage of rapamycin was adjusted weekly in accordance with average weight of mice in each treatment group. Serum samples were assayed for rapamycin concentrations by liquid chromatography-tandem mass spectrometry using a modification of Koal *et al* (*88*). Calibration was performed with matched human serum and whole blood samples, and calibration curves were linear between 0.24 and 4.98 µg/L, with a mean r value of 0.9964 (n=6), quality controls were within 10% of target concentrations and had coefficients of variation of 6.19% at 0.36 µg/L, 3.42% at 1.54 µg/L and 8.89% at 2.64 µg/L.

Mice were culled 3 weeks after the final vaccination and spleen was collected to perform the following assays.

#### Mouse IFNγ ELISpot

Mouse IFNγ ELISpot assay was performed on red blood cell–depleted splenocytes from immunized mice, which were stimulated with different peptide pools for 36 hours at 37°C, essentially as we described previously (*89, 90*). Briefly, multiscreen-IP HTS plates (Merck Millipore) were coated with anti-mouse IFNγ (clone AN18, Mabtech), and secreted IFNγ was detected with anti-mouse IFNγ biotin (clone R4-6A2, Mabtech) followed by streptavidin–alkaline phosphatase (AP) (Sigma-Aldrich) and SigmaFast BCIP (5-bromo-4-chloro-3-indolyl phosphate)/NBT (nitroblue tetrazolium; Sigma-Aldrich). Spike and RBD specific immune responses as we previously described. Spots were enumerated automatically using an ELISpot reader (AID Germany). The specific spot-forming units (SFU) per 10^6^ cells were determined by subtracting the number of spots in the unstimulated splenocytes from the number of spots in the peptide-stimulated splenocytes.

#### Mouse AIM and ICS

To assess the intracellular cytokines production, 10^6^ splenocytes were stimulated at 37◦C for 12 hrs with 4 ug/ml of SARS-CoV-2 peptide pools as described above, media or PHA served as the negative and positive controls. Brefeldin A was added for the final 4 hrs of incubation. Cells were harvested and stained with the anti-mouse surface markers CD3-BV480 (clone 17A2, 1 in 100, Cat No.565642), CD4-BV510 (clone RM4-5, 1 in 100, Cat No.563106), CD8-FITC (clone 53-6.7, 1 in 100, Cat No.553031), CD44-BV711 (clone IM7, 1 in 1000, Cat No.563971), CD62L-APC-Cy7 (clone MEL-14, 1 in 100, Cat No. 104428) antibodies, and fixable ViaDye Red (Cytek Biosciences). Cells were then fixed with 4% paraformaldehyde and permeabilized the cell membrane following intracellularly cytokine staining with anti-mouse IFN-g-APC (clone XMG1.2, Cat No.17-7311-82) and TNF-a-PE-Cy7 (clone MP6-XT22, Cat No.25-7321-82), both purchased from eBioscience.

For detection of Spike-specific T cells, freshly isolated splenocytes were cultured and stimulated with SARS-COV-2 peptides pools and control as previously described for 18 hours in a 37◦C incubator. Following the incubation, cells were stained with a multicolour antibody panel: ViadyeRed, CD3-BV480 (clone 17A2, 1 in 100, Cat No.565642), CD4-BV510 (clone RM4-5, 1 in 100, Cat No.563106), CD8-FITC (clone 53-6.7, 1 in 100, Cat No.553031), CD44-BV711 (clone IM7, 1 in 1000, Cat No.563971), CD62L-APC (clone MEL-14, 1 in 100, Cat No.55315), CD25-BV421 (clone 3C7, 1 in 100, Cat No. 566228), CD69-BV786 (clone H1.2F3, 1 in 100, Cat No. 564683), OX40-APC-Fire-750 (clone OX-86, 1 in 100, Cat No.119423). After staining, all samples were fixed with 4% paraformaldehyde and kept in dark at 4◦C fridge until performing the analysis. Unless stated all the antibodies were purchased from a commercial company (BioLegend or BD). All samples were run on a spectral flow cytometer (Cytek Aurora) and the result was analyzed using FCS express version 6.

### Statistical analysis

All statistical analyses were performed using GraphPad Prism 9.0.0 (San Diego, CA, US) or Stata Statistical Software: Release 14.2 (StataCorp, College Station, TX, US). For human observational studies, no assumptions were made about the distribution of the data sets; non-parametric tests were used in all cases for comparisons. Accordingly, Mann-Whitney and Kruskal-Wallis tests with Dunn’s correction were applied to pair-wise and multiple comparisons, respectively. For analyses of mouse experiments, one-way ANOVA with Holm’s correction was used for multiple comparison between means.

Spearman’s correlation was used to calculate all correlation coefficients. All p-values were corrected using a false discovery rate of 5%.

#### Randomized, controlled trial

Baseline characteristics and demographic data were reported as mean ± SD for normally distributed data and median ±IQR for non-normally distributed data, with categorical variables reported as frequencies. The primary analysis was intention-to-treat, with participants developing positive SARS-CoV-2 infection during the study excluded to avoid confounding. The primary outcome was the proportion of patients reaching the protective neutralization titer, analyzed using the chi-square test. Unadjusted and adjusted relative risks were calculated using the log-binomial regression model, with fixed-effects of initial immune response stratified by initial anti-RBD level and random effects of study site. Secondary outcomes were analyzed using chi-squared test for proportions and student’s t-test for comparison of normally distributed continuous data. Difference in the median value of T cell responses as measured by IFNγ ELISpot between groups was analyzed using quantile regression. Analyses were performed using Stata Statistical Software: Release 14.2 (StataCorp, College Station, TX, US).

## Data Availability

All data produced in the present work are contained in the manuscript or available upon reasonable request to the authors.

## Acknowledgments

We thank everyone who contributed to this study, particularly those who participated in the trials during the height of the pandemic. We would also like to thank the staff of the Royal Adelaide Hospital Medical Day Unit and Specialist Vaccination Clinic, and acknowledge Dennis Penglis and Tina Petrou for their assistance with serological studies.

## Author contributions

Conceptualization: GBP, MJT, PH, SCB, SJC, BGB, PTC.

Methodology: GBP, MJT, CSC, TS, JS, CMH, MGM, ZAM, PGV, ST, TY, SJC, BGB, PTC.

Formal analysis: GBP, MJT, CSC, BS, TY.

Investigation: GBP, MJT, CSC, TS, AELY, JS, CMH, BS, MGM, ZAM, PGV, SK, JKJ, BZS, IC, SMS, BCS, AAg, VM, AAk, PRH, TY.

Visualization: GBP, MJT, CSC, BS, GB.

Funding acquisition: PH, SCB, SJC, BGB, PTC.

Project administration: GBP, MJT, CSC, TS, JS, CJD, SJC, BGB, PTC.

Supervision: PRH, CMH, PH, SCB, SJC, BGB, PTC.

Writing – original draft: GBP, MJT.

Writing – review & editing: All authors reviewed the manuscript.

## Data and materials availability

All data are available in the main text or the supplementary materials.

## SUPPLEMENTARY MATERIALS

**Fig. S1 (relates to Figs. 2 & 3).**
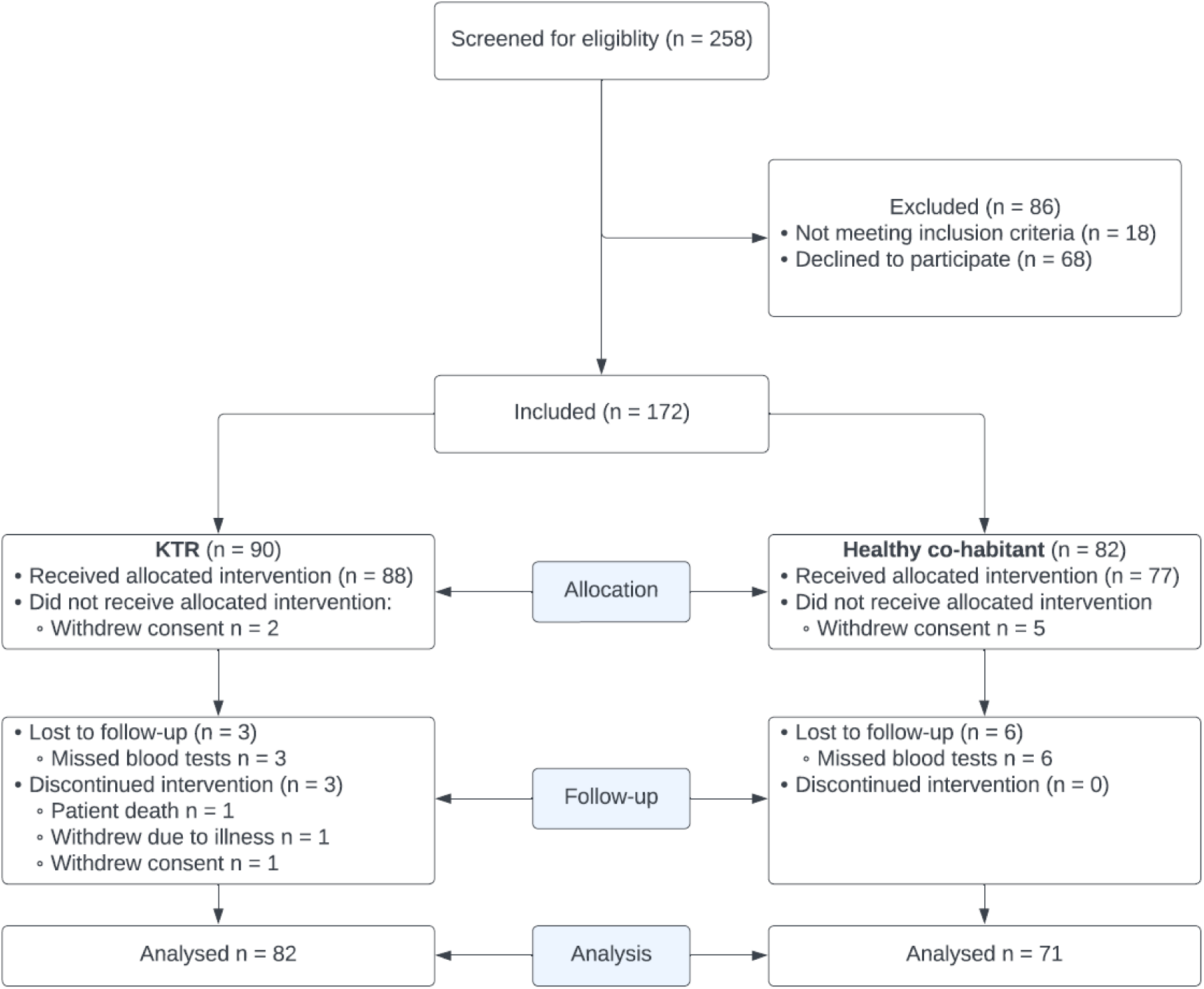
CONSORT diagram for the REVAX observational study.

**Fig. S2 (relates to Fig. 2).**
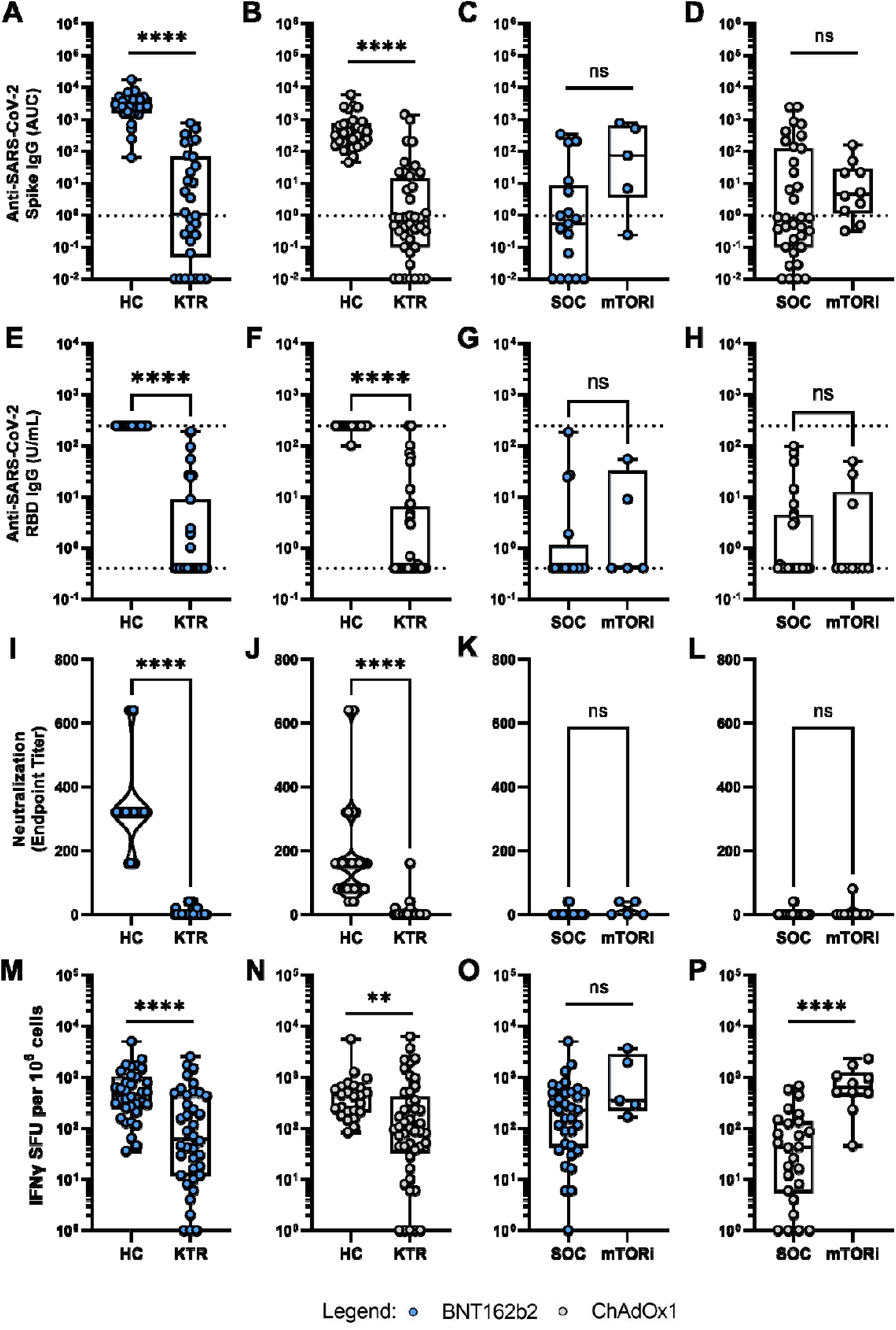
Immune response of kidney transplant recipients (KTRs) to a primary COVID-19 vaccination schedule stratified by vaccine type. Responses of KTRs (n = 83) are compared against healthy controls (HC; n = 71), and between clinically comparable calcineurin inhibitor (SOC; n = 41)- and mTOR inhibitor (mTORi; n = 15)-based three-drug immunosuppressive regimens. (**A-D**) Endpoint titers are reported as area under the curve (AUC). Dashed lines represent detection threshold (mean AUC +L2 SD of negative controls). (**E-H**) Log-scale comparison of anti-RBD IgG. The detection range for the assay is 0.4 – 250 U/mL, as demarcated by dotted lines. Percentage of seropositive individuals (above 0.4 U/mL) is shown for each group. (**I-L**) Endpoint titers (minimum serum dilution) for 50% neutralization of live SARS-CoV-2 virus (ancestral strain) entry into target cells. (**M-P**) Log-scale comparison of Spike-specific IFNγ-secreting T cell responses measured as spot-forming units (SFU) by ELISpot. **Comparisons between groups by Mann-Whitney U test (flat bars) or Fisher’s exact test (crooked bars): ns, non-significant; *p<0.05; **p<0.01; ***p<0.001; ****p<0.0001.**

**Fig. S3 (relates to Fig. 2).**
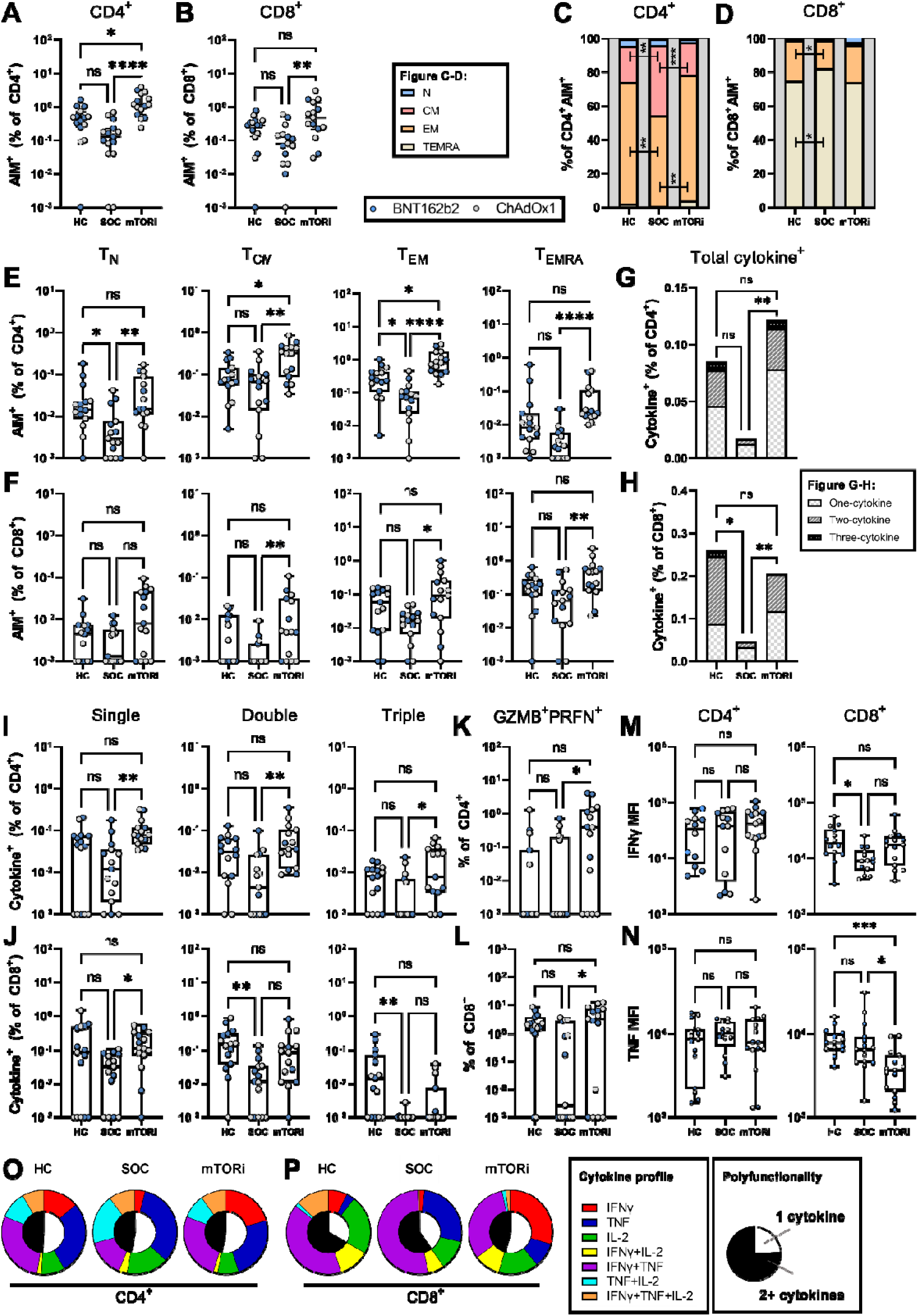
Phenotype and function of the memory T cell response: comparison of mTORi-based versus CNI-based (SOC) three-drug regimens following 2 doses of COVID-19 vaccine. (**A**) Percentage of Spike-specific (AIM^+^) cells within the CD4^+^ and CD8^+^ T cell compartments. (**C-D**) Contribution of each phenotype to the CD4^+^ and CD8^+^ AIM^+^ populations. Naïve: T_N_, CCR7^+^CD45RA^+^; Central Memory: T_CM_, CCR7^+^CD45RA^−^; Effector Memory: T_EM_, CCR7^−^CD45RA^−^; Terminally Differentiated Effector Memory: T_EMRA,_ CCR7^−^CD45RA^+^. (**E-F**) Frequency of each AIM^+^ subpopulation as a percentage of the total CD4^+^ or CD8^+^ T cell populations. (**G-H**) Percentage of CD4^+^CD154^+^ and CD8^+^ T cells upregulating at-least one cytokine in response to stimulation. (**I-J**) Log-scale comparison of the percentage of CD4^+^ and CD8^+^ T cells single, double, or triple positive for any combination of IL-2, TNF and IFNγ cytokines. (**K-L**) Log-scale comparison of the percentage of CD4^+^ and CD8^+^ T cells positive for intracellular granzyme B (GZMB^+^) and perforin (PRFN^+^) following stimulation with Spike peptides. (**M-N**) IFNγ and TNF mean fluorescence intensity (MFI) of Spike-specific CD4^+^ and CD8^+^ T cells. (O-P) Median cytokine expression profile (outer pie chart) and proportion of cytokine positive cells that are polyfunctional (inner pie chart) of CD4^+^ and CD8^+^ T cells. **Statistical analysis by Kruskal-Wallis with Dunn’s correction: ns, non-significant; *p<0.05; **p<0.01; ***p<0.001; ****p<0.0001.**

**Fig. S4 (relates to Fig. 2).**
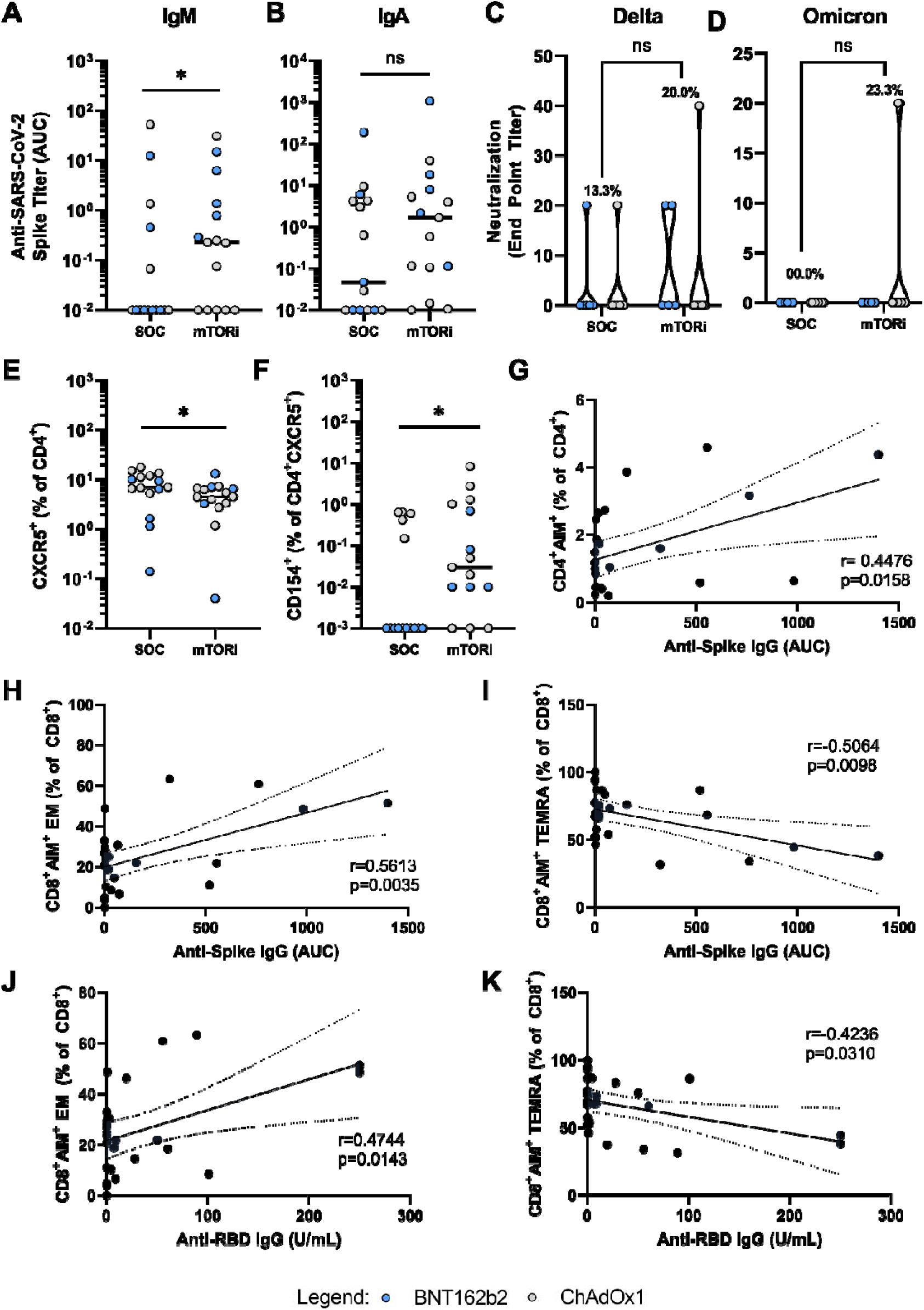
**Characteristics and correlates of antibody response** to primary COVID-19 vaccination in kidney transplant recipients receiving an mTORi-based three-drug immunosuppression regimen (n = 15) compared to the standard-of-care regimen (n = 15). **(A-B)** Log-scale comparison of anti-Spike IgM and IgA. Titers were assessed for eight serum dilutions and are presented as area under the curve (AUC). **(C-D)** Log comparison of live virus neutralization endpoint titer-minimum serum dilution for 50% neutralization of live SARS-CoV-2 virus (Delta and Omicron BA.1) entry into target cells. **(E-F)** Frequency of total and Spike-specific circulating T follicular helper (cT_FH_) cells, measured as percentage CXCR5^+^ of CD4^+^ T cells and percentage of CD154^+^ of CD4^+^CXCR5^+^ population following stimulation with Spike peptides, respectively. **(G–K)** Correlates of anti-Spike and anti-RBD IgG titers in KTRs receiving mTORi-based triple therapy. **Analysis by Mann-Whitney *U* test (straight bars) or Fisher’s exact test (crooked bars), and Pearson’s correlation: ns, non-significant; *p<0.05; **p<0.01; ***p<0.001; ****p<0.0001.**

**Fig. S5 (relates to Fig. 3).**
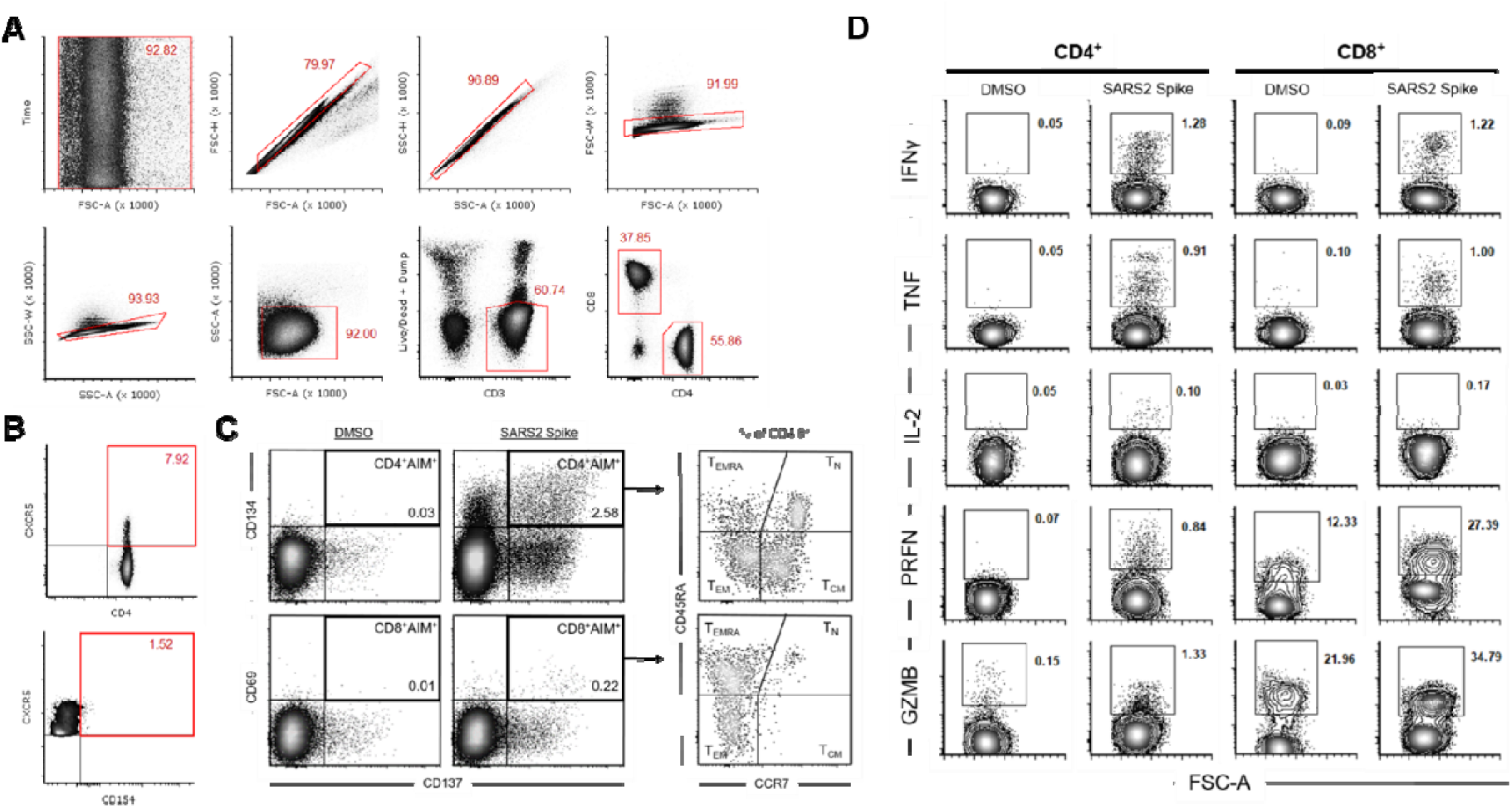
Gating strategy for analysis of activation-induced marker (AIM) expression and intracellular cytokine staining (ICS). (**A**) Singlets were identified and T cells defined as CD3^+^ at the exclusion of CD14^+^CD19^+^ cells and dead cells (dump gate). (**B**) cT_FH_ cells were defined as CD4^+^CXCR5^+^, and Spike-specific cT_FH_ cells quantified based on expression of CD154 (CD40L) following antigen stimulation. (**C**) Spike-specific memory T cell were defined as CD4^+^CD134^+^CD137^+^ or CD8^+^CD69^+^CD137^+^ following antigen stimulation. (**D**) Effector molecule producing cells were quantified within the CD4^+^ and CD8^+^ gates. Cytokine-producing antigen-specific CD4^+^ T cells were defined by co-expression of CD154.

**Fig. S6 (relates to Fig. 3).**
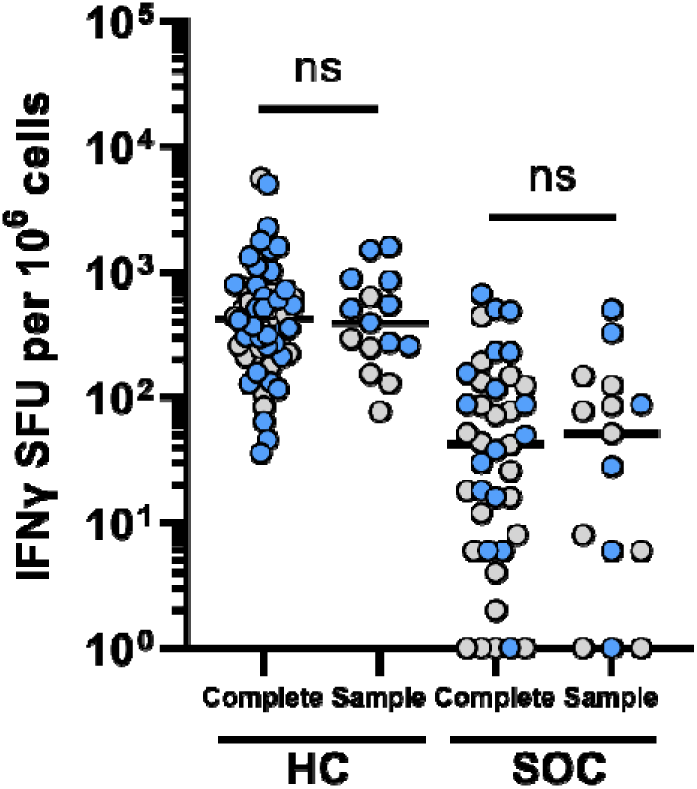
**Comparison of IFNγ ELISpot response between the complete cohort (Fig. 2) and sample populations (Fig. 3, figs. S3-S4)** following 2 doses of COVID-19 vaccine. Isolated peripheral blood mononuclear cells (PBMC) were stimulated for 18 hours with spike glycoprotein-derived peptides and the frequency of IFNγ-secreting cells measured as spot-forming units (SFU) by ELISpot. **Comparisons between groups by Mann-Whitney *U* test: ns, non-significant; *p<0.05; **p<0.01; ***p<0.001; ****p<0.0001.**

**Fig. S7 (relates to Fig. 3).**
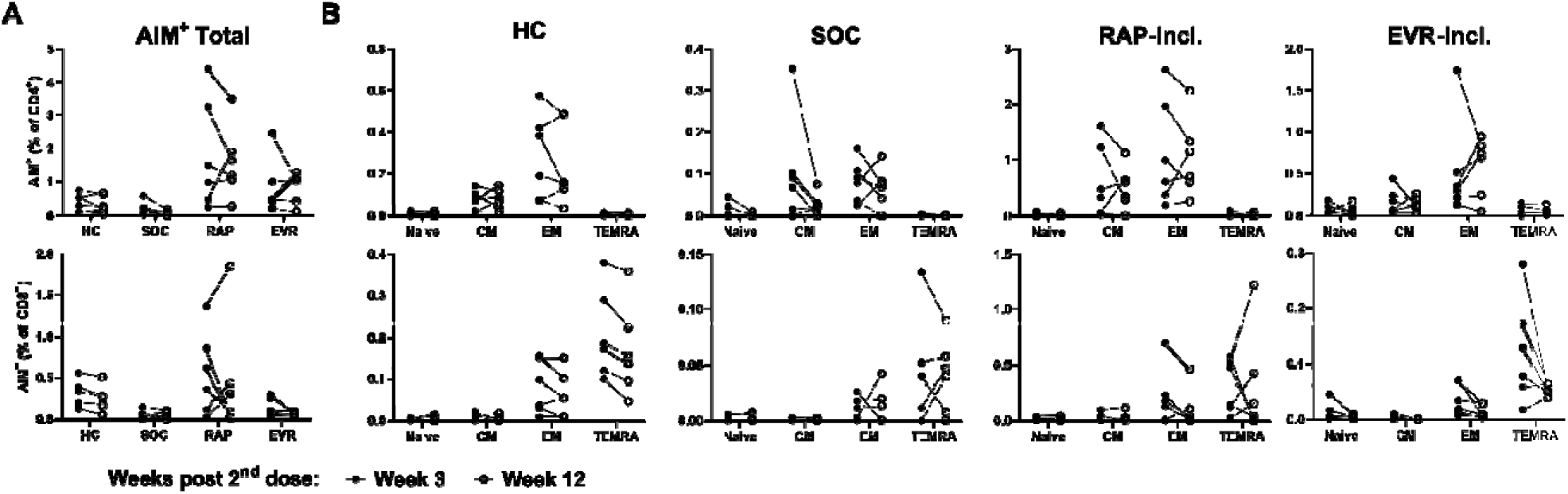
**Follow up analysis of Spike-specific T cell responses by AIM assay at 3 weeks versus 12 weeks** post second COVID-19 vaccine dose in KTRs on CNI-based SOC (n = 6), rapamycin-inclusive (n = 6) or everolimus-inclusive (n = 6) therapies, and healthy controls (n = 6). **(A)** Frequency of AIM^+^CD4^+^ and AIM^+^CD8^+^ T cells. **(B)** Frequency of AIM^+^ naïve (T_N_, CCR7^+^CD45RA^+^), central memory (T_CM_, CCR7^+^CD45RA^−^), effector memory (T_EM_, CCR7^−^CD45RA^−^) and terminally differentiated effector memory (T_EMRA,_ CCR7^−^CD45RA^+^) cells as a percentage of total CD4^+^ and CD8^+^ T cell compartments.

**Fig. S8 (relates to Figure 3).**
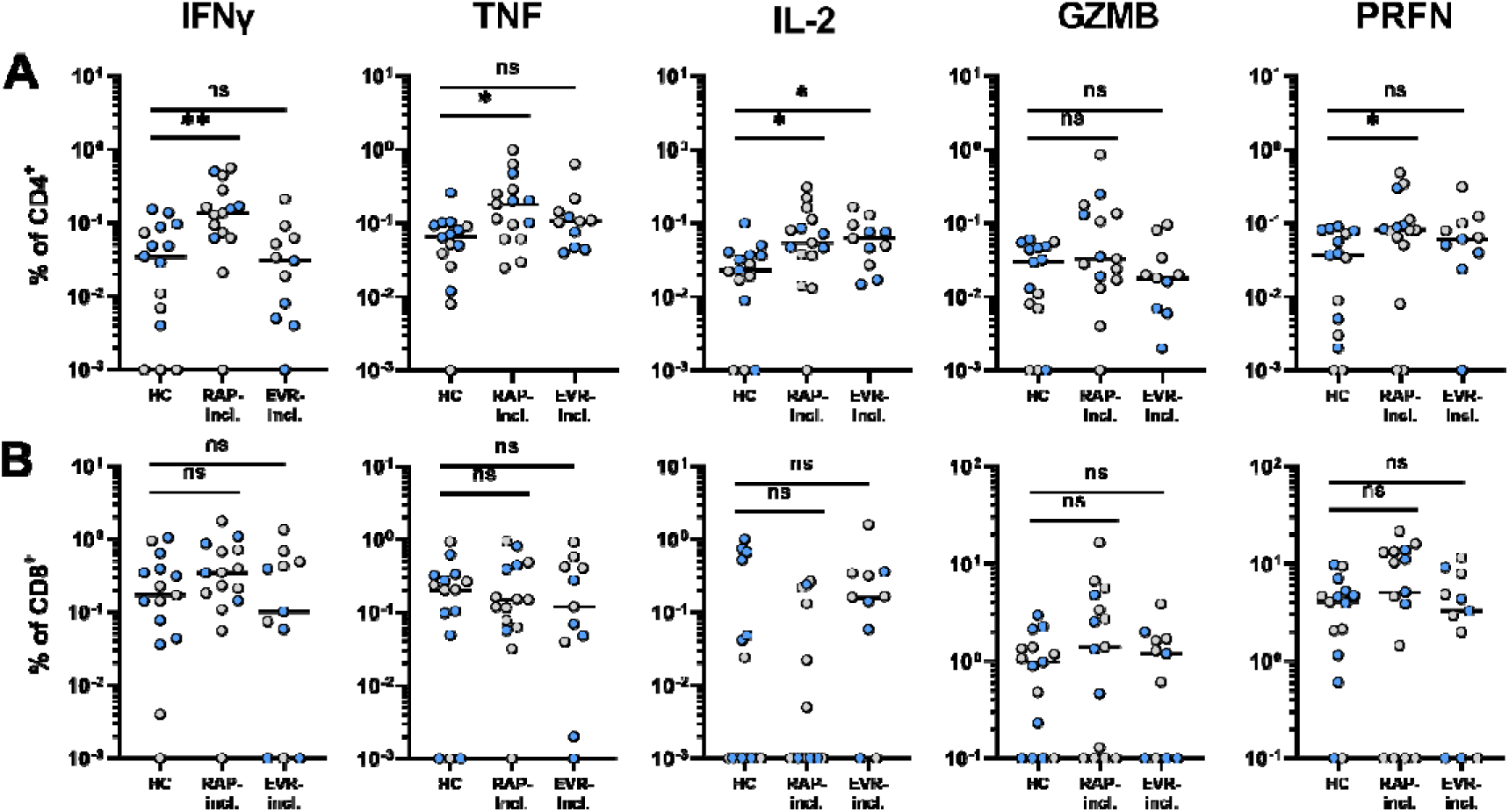
**Effector profile of Spike-specific T cells** in KTRs on rapamycin-inclusive (RAP-incl; n = 14) and everolimus-inclusive (EVR-incl; n = 11) immunosuppressive therapies, and healthy controls (HC; n = 15) following 2 doses of COVID-19 vaccine. **(A, B)** Effector molecule expression by CD4^+^CD154^+^ and CD8^+^ T cells in response to Spike peptides. **Statistical analysis by Kruskal-Wallis with Dunn’s correction comparing each group to healthy controls: ns, non-significant; *p<0.05; **p<0.01; ***p<0.001; ****p<0.0001.**

**Fig. S9.**
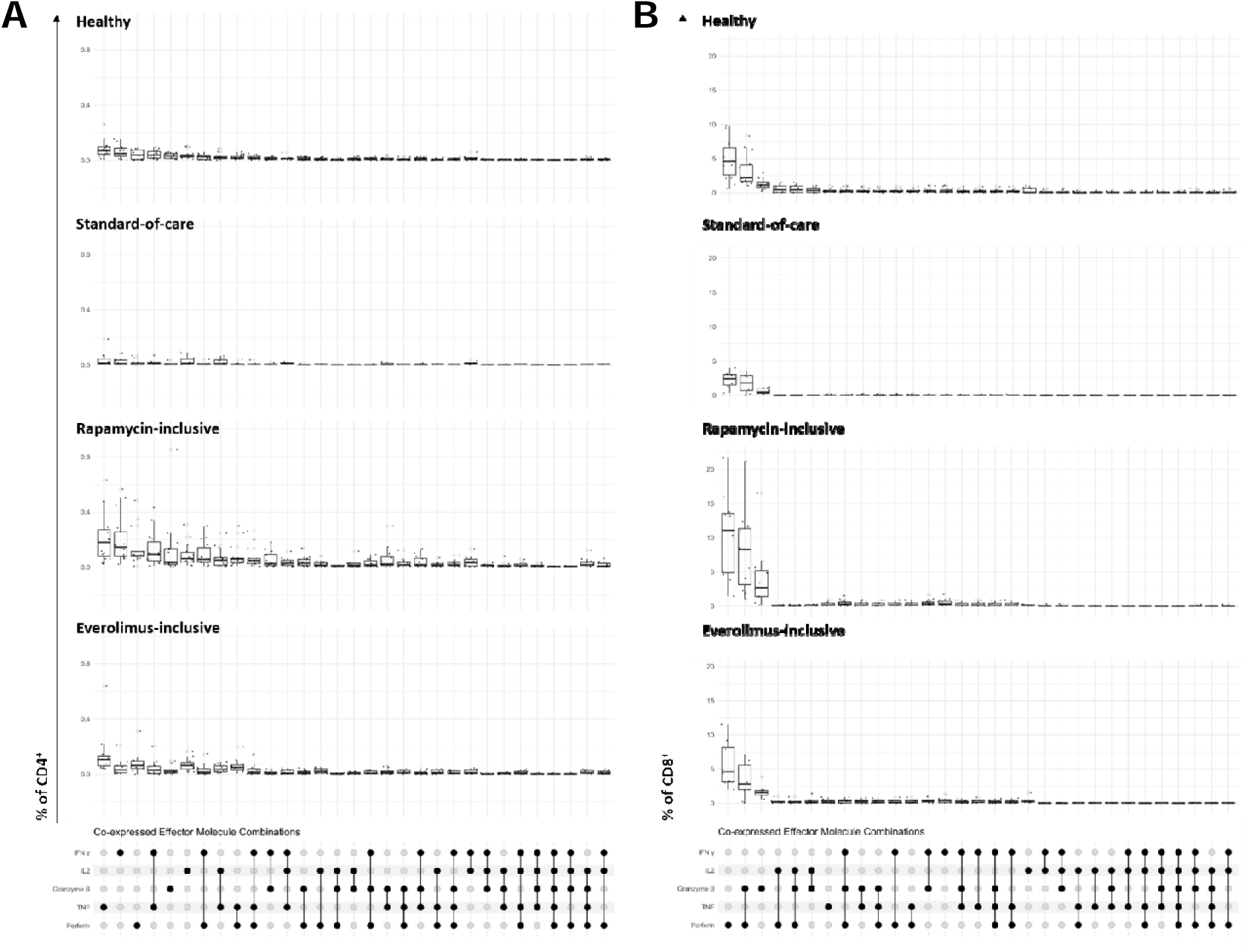
**Effector molecule co-expression combinations for CD4^+^ and CD8^+^ T cells** in response to stimulation with SARS-CoV-2 Spike peptides. Combinations are ordered by high to low frequency in the healthy group for CD4^+^ **(A)** and CD8^+^ **(B)** T cells.

**Fig. S10 (relates to Fig. 4).**
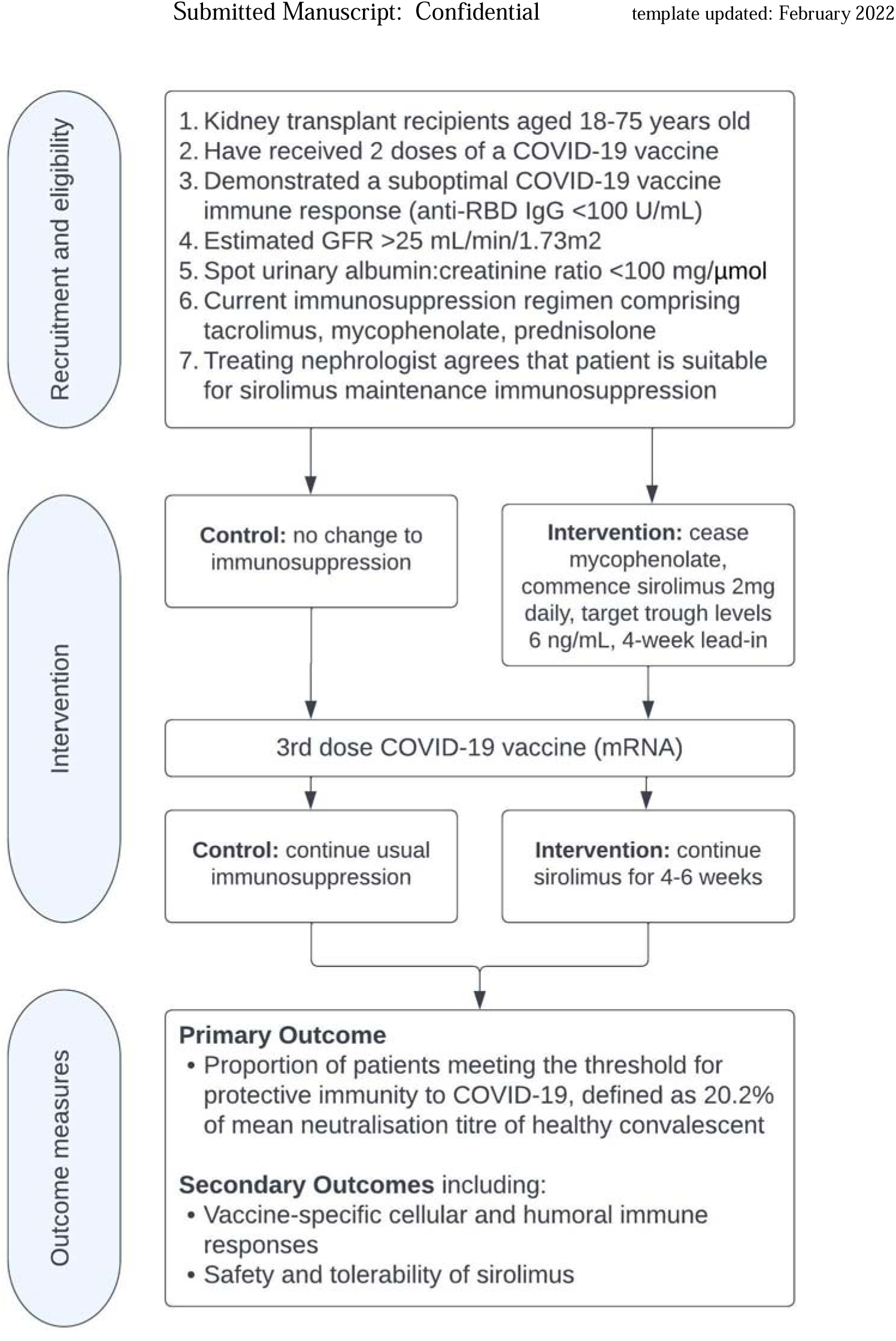
**RIVASTIM-Rapamycin overview diagram** outlining inclusion/exclusion criteria and study design (adapted from Tunbridge et al. 2022 (*55*)).

**Fig. S11 (relates to Fig. 4).**
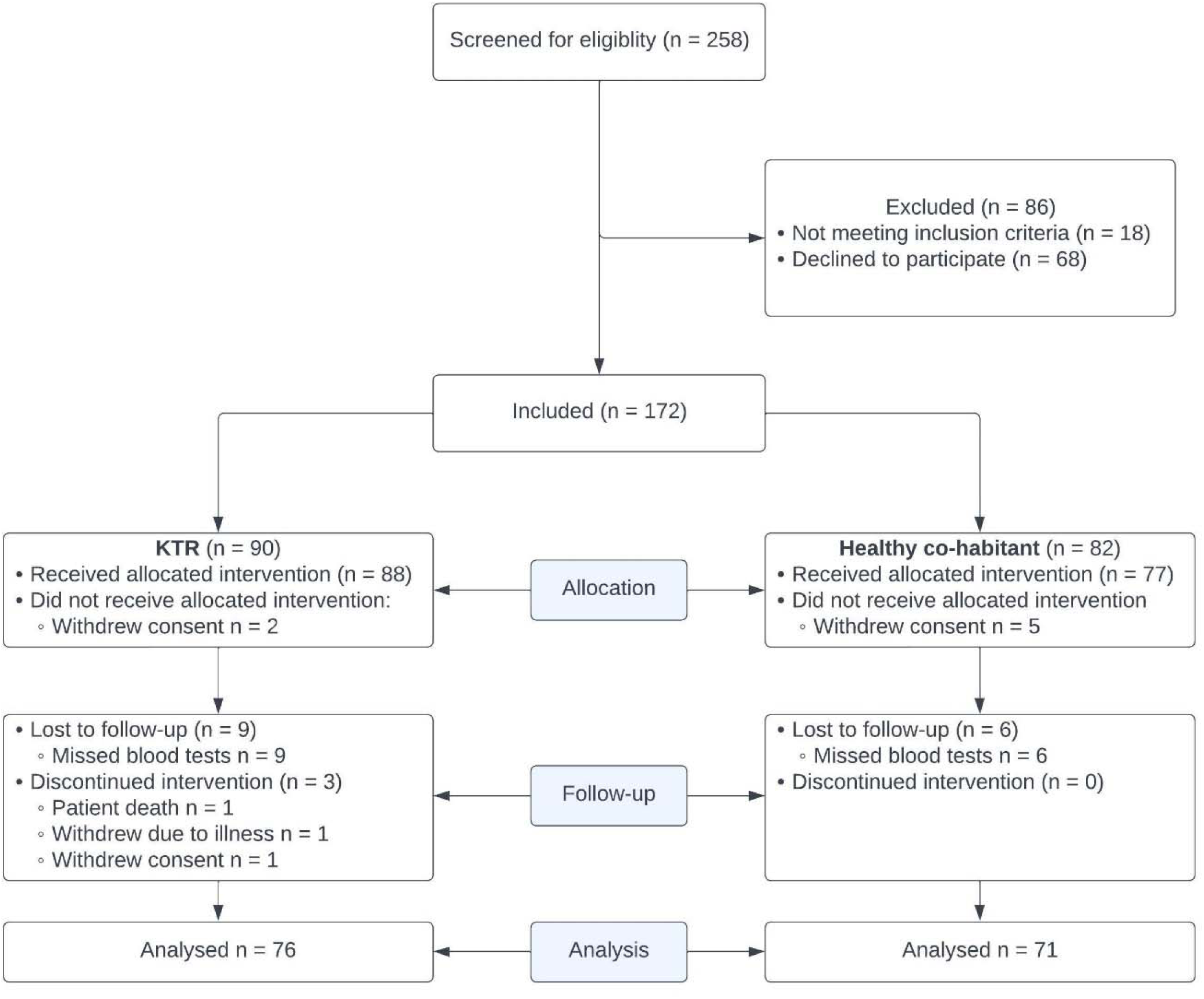
RIVASTIM-Rapamycin PRISMA diagram.

**Table S1 (relates to Fig. 4).** RIVASTIM-Rapamycin safety reporting.

|  | Rapamycin switch (n = 25) | No switch (n = 23) | P-value |
| --- | --- | --- | --- |
| Patients with any AEs | 7 (28) | 2 (9) | 0.09 |
| Patients with severe AEs | 2 (8) | 1 (4) | 0.60 |
| Patients with AE leading to death | 0 (0) | 0 (0) |  |
| AEs* | 7 | 3 |  |
| • Infection | 2 | 1 | 0.58 |
| • Renal/urinary | 2 | 0 | 0.16 |
| • Cardiac | 1 | 0 | 0.32 |
| • Gastrointestinal | 1 | 0 | 0.32 |
| • Hepatobiliary | 0 | 1 | 0.30 |
| • Nervous system | 0 | 1 | 0.30 |
| • Surgical procedures | 1 | 0 | 0.32 |
| AE attribution | 7 | 3 |  |
| • Neither the vaccine nor the treatment group | 7 | 2 |  |
| • Receiving the vaccine (3rd dose) | 0 | 1 |  |
| • Intervention (Rapamycin switch) | 0 | 0 |  |
| • Control (no switch) | 0 | 0 |  |
| • Both the vaccine AND the intervention/control | 0 | 0 |  |

**Table S2 (relates to Fig. 4).** Adverse events following immunisation (AEFI) assessed throughout RIVASTIM-Rapamycin study period up to 4-6 weeks post-vaccination.

| Factor | Arm | N | Mean change<br>(Final visit – baseline) | SD | P-value |
| --- | --- | --- | --- | --- | --- |
| Serum creatinine | Rapamycin switch | 25 | 0.0 | 2.5 |  |
|  | No switch | 23 | 4.0 | 3.7 |  |
|  | Difference between 2 arms |  | -4.0 | 4.7 | 0.39 |
| eGFR | Rapamycin switch | 25 | 0.3 | 2.1 |  |
|  | No switch | 23 | -1.9 | 1.0 |  |
|  | Difference between 2 arms |  | 2.2 | 2.4 | 0.35 |
| Urine ACR | Rapamycin switch | 24 | 15.1 | 7.1 |  |
|  | No switch | 22 | -1.7 | 2.5 |  |
|  | Difference between 2 arms |  | 16.8 | 7.8 | **0.038** |

**Table S3 (relates to Fig. 4).** Influence of existing immunity on antibody response to booster vaccination. Proportion of participants in each trial arm that reached a threshold of serological anti-SARS-CoV-2 RBD IgG titer ≥ 100 U/mL. No significant association was found by chi-square test of independence χ^2^ (1, N = 45) = 0.03, p = 0.87.

|  | Rapamycin<br>Switch | No switch | p-value |
| --- | --- | --- | --- |
|  | N=25 | N=23 |  |
| **Anti-RBD IgG at 8 week** |  |  | 0.82 |
| <0.4 units/mL | 1 (4) | 0 (0) |  |
| 0.4~100 units/mL | 11 (44) | 10 (43) |  |
| $\geq 100$ units/mL | 12 (48) | 11 (48) | |
| Missing | 1 (4) | 2 (9) |  |
| **Baseline Anti-RBD IgG < 0.4 units/mL** | **9** | **7** |  |
| Anti-RBD IgG at 8 week |  |  | 0.28 |
| <0.4 units/mL | 1 (11) | 0 (0) |  |
| 0.4~100 units/mL | 6 (67) | 6 (86) |  |
| $\geq 100$ units/mL | 2 (22) | 0 (0) | |
| Missing | 0 (0) | 1 (14) |  |
| **Baseline Anti-RBD IgG $\geq 0.4$ units/mL)** | **16** | **16** |  |
| Anti-RBD IgG at 8 week |  |  | 0.79 |
| 0.4~100 units/mL | 5 (31) | 4 (25) |  |
| $\geq 100$ units/mL | 10 (62) | 11 (69) | |
| Missing | 1 (6) | 1 (6) |  |

**Fig. S12 (relates to Fig. 4).**
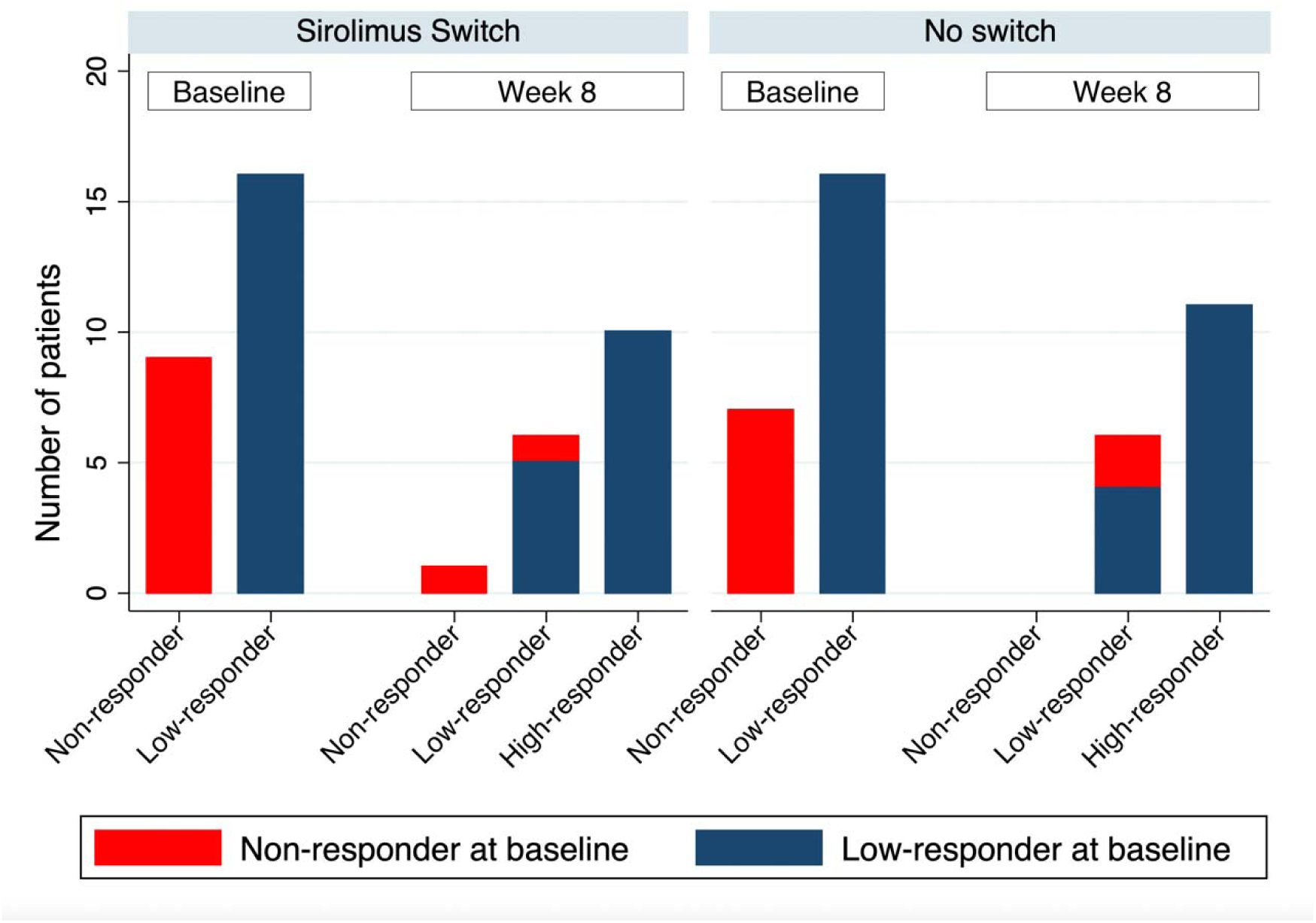
Outcome of block randomization. Number of patients in each response category by study arm and study visit. Response categories defined as non-responder (anti-RBD IgG <0.4 U/mL), low-responder (anti-RBD IgG 0.4-100 U/mL), and high-responder (anti-RBD IgG >100 U/mL). No significant difference in proportions was seen between switch and no-switch groups.

**Table S4 (relates to Fig. 4).** Development of COVID-19 following randomization. A chi-square test of independence showed no significant association, χ^2^ (1, N = 51) = 2.62, p = 0.11.

| Development of COVID-19<br>following randomization |  | <u>Randomization arm</u> |  |  |
| --- | --- | --- | --- | --- |
|  |  | Rapamycin<br>Switch | No switch | Total |
|  | No | 25 | 23 | 48 |
|  | Yes | 3 | 0 | 3 |
|  | Total | 28 | 23 | 51 |

**Fig. S13 (relates to Fig 5).**
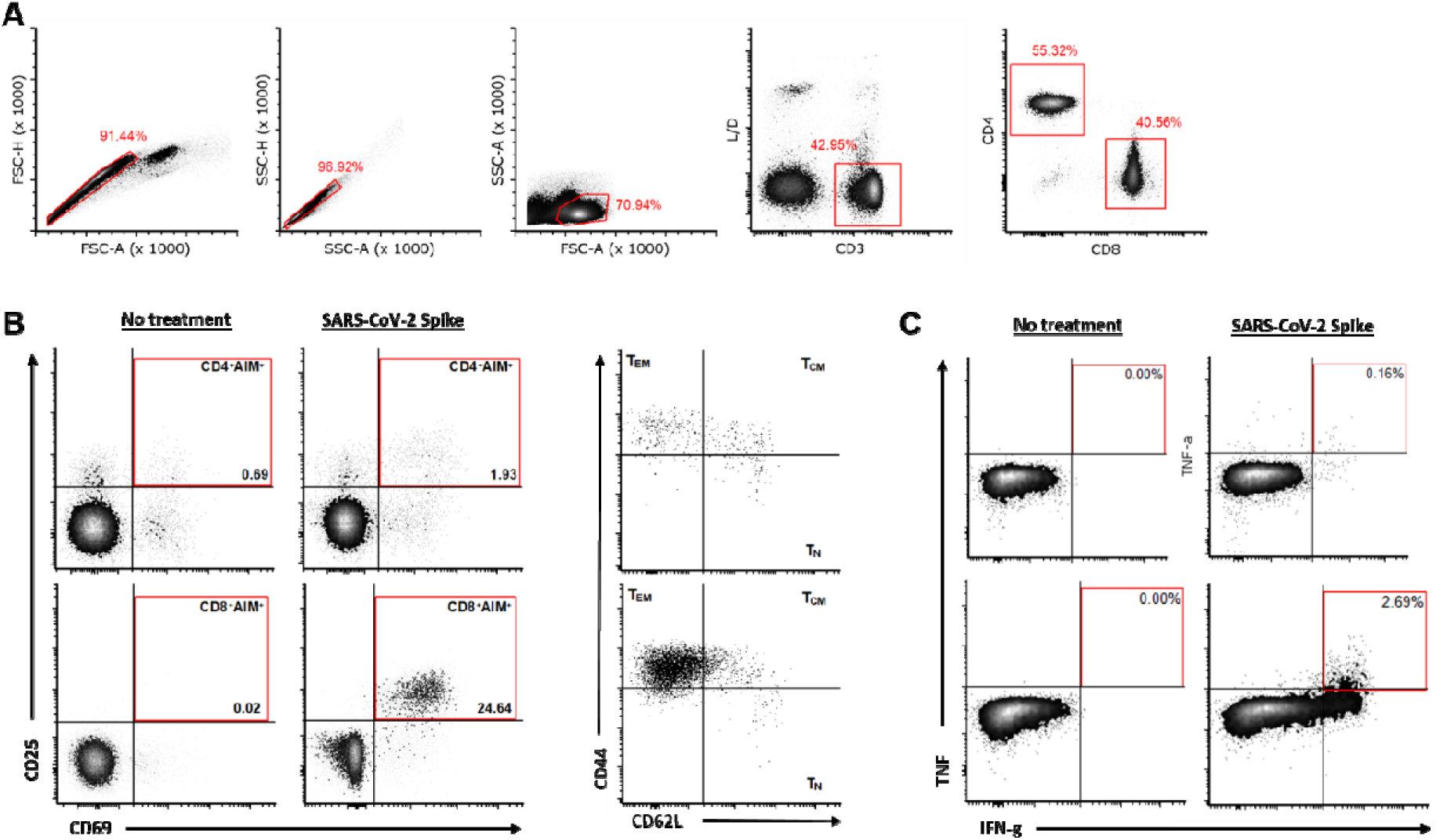
Gating strategy for analysis of activation-induced marker (AIM) expression and intracellular cytokine staining (ICS) by mouse splenocytes. (**A**) Doublet exclusion and gating of viable CD3^+^CD4^+^ and CD3^+^CD8^+^ T cells. (**B**) Spike-specific memory T cells were defined by co-expression of CD25 and CD69 following antigen stimulation. Memory phenotype of AIM^+^ cells was assessed based on expression of CD44 and CD62L. (**C**) IFNγ and TNF producing cells were quantified within the CD4^+^ and CD8^+^ gates.

**Fig. S14 (relates to Fig 5).**
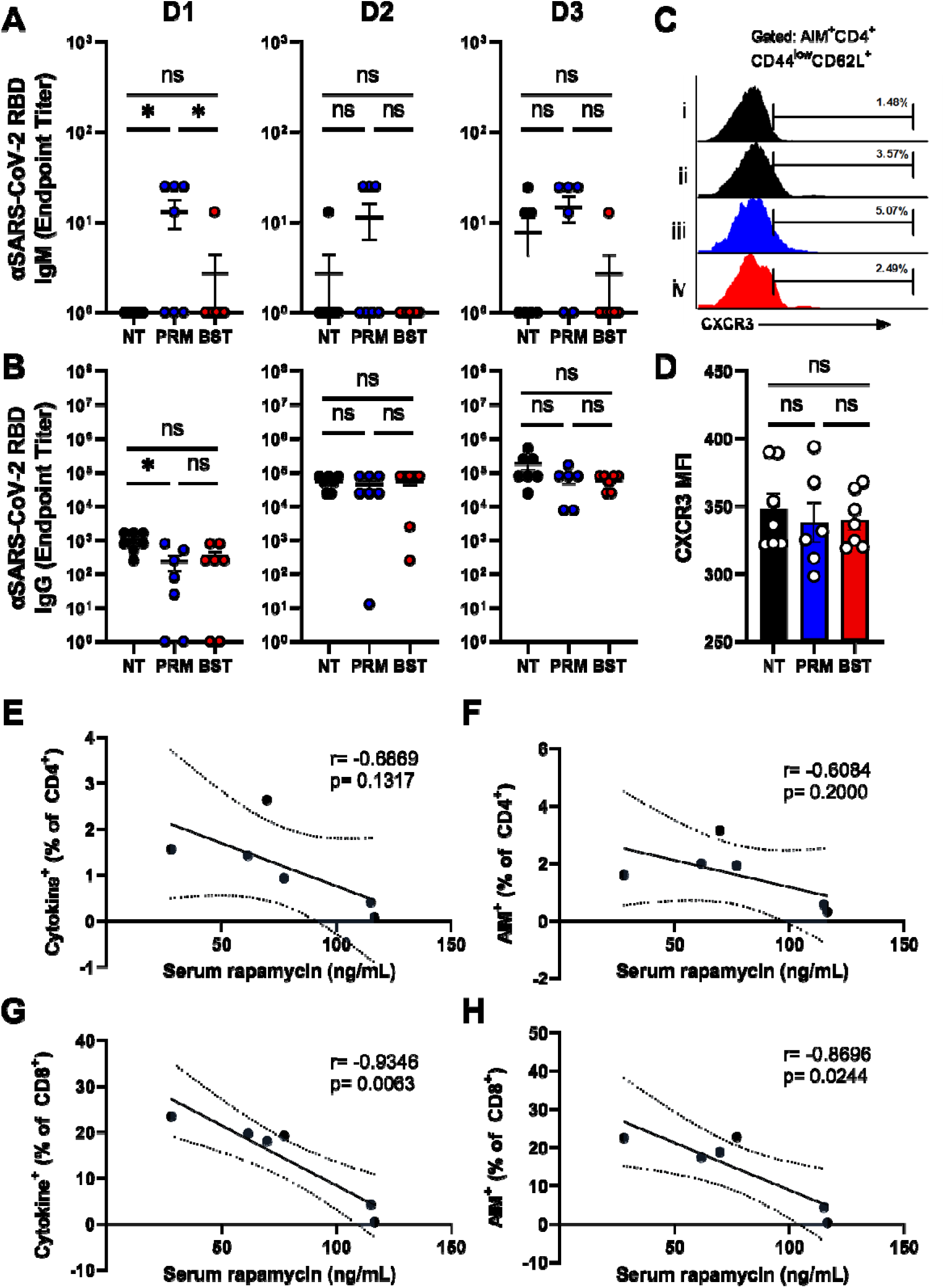
Effect of rapamycin on anti-RBD titer and CXCR3 expression by putative T_SCM_. (**A-B**) Serum titers of anti-RBD IgM (**A**) and IgG (**B**) following the first (D1), second (D2) and third (D3) vaccine doses. (**C-D**) Percentage (**C**) and mean fluorescence intensity (**D**) of CXCR3 expression by AIM^-^ naïve-like (CD44^low^CD62L^+^) CD4^+^ T cells (i) and AIM^+^ naïve-like (CD44^low^CD62L^+^) T cells from no treatment (ii), pre-primary (iii), and pre-booster (iv) rapamycin treated mice. (**E-H**) Inverse correlation of Spike-specific T cell responses with rapamycin concentrations in the serum at time of T cell analysis.

## SUPPLEMENTARY RESULTS

The main text of this article focuses on comparing KTRs on mTORi to healthy controls, with the aim of demonstrating that mTORC1 inhibition has a positive effect on T cell memory formation in humans. Here, we compare in greater detail the difference between the CNI-based standard-of-care (SOC) versus mTORi-based three-drug immunosuppression regimens on the antigen-specific T cell response to primary COVID-19 vaccination.

### Correlates of antibody response in patients on mTORi-based protocols

The higher titer of anti-Spike IgG in the mTORi group relative to the SOC group was surprising given the important role of mTORC1 signaling in the formation of T follicular (T_FH_) cells and germinal centers in mice (*26*). Consistent with such a role, patients receiving the mTORi-based protocol demonstrated a reduced frequency of circulating (c)T_FH_ cells (defined as CD4^+^CXCR5^+^ T cells) compared with the SOC group (4.57% vs 7.09% CXCR5^+^ of CD4^+^, p=0.03670) (fig. S3). However, a greater proportion of cT_FH_ cells in this group upregulated CD154 in response to stimulation with Spike peptides, suggesting that the reduced cT_FH_ cell frequency may be offset by a greater proportion of antigen-specific cells (0.030% vs 0.001% CD154^+^ of CD4^+^CXCR5^+^ T cells, p=0.0408) (fig. S3). While antigen-specific cT_FH_ cell frequency has been associated with anti-Spike IgG titer in healthy individuals and kidney transplant recipients (*5*), CD154^+^CD4^+^CXCR5^+^ T cells did not correlate with IgG response in KTRs on mTORi, and therefore is unlikely to account for the higher IgG titers in this group compared to the SOC protocol. Additional correlation analysis identified moderate positive correlations of anti-Spike IgG with total AIM^+^ CD4^+^ T cells, and with the proportion of AIM^+^ CD8^+^ T cells that were of an effector memory phenotype (T_EM_) (fig. S3). Reciprocally, the proportion of AIM^+^ CD8^+^ that were of a T_EMRA_ phenotype was negatively correlated with anti-Spike IgG (fig. S3).

### Impairment of T cell phenotype and function following primary COVID-19 vaccination in KTRs on standard-of-care CNI-based but not mTORi-based three-drug protocols

In contrast to patients on the mTORi three-drug protocol, those on SOC demonstrated significant impairment of the Spike-specific T cell response relative to healthy controls (Fig 2, fig. S4). This deficit was most pronounced in the >10-fold reduction in IFNγ-secreting Spike-specific T cells by ELISpot (Fig. 2), an observation that reflected both a reduced frequency of IFNγ-producing CD8^+^ T cells, and a reduction in the amount of IFNγ produced per cell (as measured by MFI in IFNγ^+^ cells, fig. S4).

This impairment in frequency of Spike-specific T cells was most pronounced in the T_EM_ compartment, resulting in a skewed phenotype of Spike-specific T cell memory in KTRs receiving the SOC CNI-based protocol relative to healthy individuals, which was not present in the mTORi group (fig. S4). CD4^+^ T cells in patients on SOC therapy were preferentially of a T_CM_ rather than T_EM_ phenotype relative to healthy controls (fig. S4), and a similar bias was observed in the CD8^+^ T cell compartment, with Spike-specific memory cells forming of a T_EMRA_ rather than T_EM_ phenotype (fig. S4).

Direct comparison of KTRs on the two clinically comparable protocols found that the mTORi-based protocol was associated with greater frequencies of all CD4^+^ and CD8^+^ T cell memory populations (fig. S4), as well as greater frequencies of cytokine producing CD4^+^ and CD8^+^ T cells and stronger cytotoxic recall (GZMB^+^PRFN^+^) responses (fig. S4).

